# Strategies to Achieve HIV and HCV Infection Incidence Targets Among People Who Inject Drugs: A Stochastic Network-Based Multi-Disease Transmission Modeling Study

**DOI:** 10.1101/2025.05.22.25328020

**Authors:** Marissa B. Reitsma, Lin Zhu, Hasan Symum, Nathan W. Furukawa, Dita Broz, Kevin P. Delaney, Eliza R. Ennis, Angela T. Estadt, Senad Handanagić, Dafna Kanny, Benjamin P. Linas, Nisha Nataraj, Douglas K. Owens, Teresa Puente, Liisa M. Randall, Jeremy D. Goldhaber-Fiebert, Joshua A. Salomon, the NHBS Study Group

**Author notes:** Corresponding author: Marissa B. Reitsma, PhD Department of Health Policy, Stanford University Address: Encina Commons, 615 Crothers Way, Stanford, CA 94305.

## Abstract

**Background:** The United States aims to reduce the incidence of human immunodeficiency virus (HIV) and hepatitis C virus (HCV) infections by 90% by 2030.

**Objective:** To identify strategies for achieving incidence reduction goals by scaling interventions that address the syndemic of substance use disorder, HIV infection, and HCV infection among people who inject drugs (PWID).

**Design:** Stochastic agent-based multiplex network model.

**Setting:** Urban areas in the United States.

**Participants:** People who inject drugs.

**Interventions:** Scenarios scaled interventions from current baseline to moderate and high values. Prevention and cessation interventions were increased 15 (moderate) and 30 (high) percentage points. Test and treat interventions increased the percentage of PWID with current HCV infection achieving sustained virologic response per year from 3% to 16% (moderate) and 28% (high) and the percentage of PWID with HIV that were virally suppressed from 44% to 58% (moderate) and 71% (high).

**Measurements:** HIV and HCV infection incidence among PWID over ten years, and quality-adjusted life-years, discounted at 3% annually, over 80 years.

**Results:** High coverage across all three intervention strategies resulted in an 86% (95% Uncertainty Interval (UI): 72-96%) decrease in new HIV infections, a 90% (95% UI: 87-94%) decrease in new HCV infections, and an increase of 1.8 (95% UI: 1.6-2.0) discounted quality-adjusted life-years among PWID. Moderate coverage across all three strategies yielded 62% (95% UI: 39-81%) and 68% (95% UI: 61-74%) decreases in new HIV and HCV infections among PWID, respectively. Increasing cessation of injection consistently produced the largest gains in quality-adjusted life-years.

**Limitations:** Model did not examine specific interventions or economic costs. Parameters were not representative of all urban areas or all PWID.

**Conclusion:** Increases in survival and health-related quality of life for PWID can be achieved by scaling syndemic-focused intervention strategies.

**Primary Funding Source:** This project was funded by the Centers for Disease Control and Prevention, National Center for HIV, Viral Hepatitis, STD, and TB Prevention Epidemiologic and Economic Modeling Agreement (NEEMA; award #NU38PS004651) and the National Institute on Drug Abuse.

**Disclaimer**: The findings and conclusions in this report are those of the authors and do not necessarily represent the official position of the Centers for Disease Control and Prevention, the National Institutes of Health, or authors’ affiliated institutions.

## Background

The United States established 10-year goals to reduce the numbers of new human immunodeficiency virus (HIV) and new hepatitis C virus (HCV) infections by 90% by 2030.^1–3^ Strategies to reach these targets are outlined in coordinated national strategic plans, which call attention to the ongoing syndemic of substance use disorder, viral hepatitis, HIV, and other sexually transmitted infections (STIs) among people who inject drugs (PWID).^1,2,4^ Combatting this syndemic is key to attaining national incidence targets, because the number of PWID has increased in the past decade, and PWID face a disproportionately high burden of HIV and HCV infections.^1,2,5^ An estimated 72% of new HCV infections and 7% of new HIV infections are among PWID, whereas the estimated 3.7 million PWID represent 2% of the adult U.S. population.^1,2,5^ Strategies to reduce the burden of HIV and HCV infections among PWID include testing and treatment of infections, treating substance use disorder, and expanding prevention interventions, including syringe services programs (SSPs), behavioral interventions (e.g., consistent condom use), and pre-exposure prophylaxis (PrEP) for HIV.

As resources are mobilized to scale treatment and harm reduction strategies to end the HIV epidemic and eliminate HCV infection in the United States, model-based simulation analyses can inform the design of programs, including the levels of coverage necessary to achieve identified targets.^6–9^ Although models depend on data and assumptions invoked in their design and parameterization, they enable efficient exploration of both short-term and long-term outcomes in a transparent and controlled environment.^10–15^

Prior model-based analyses of strategies to reduce the burden of HIV and HCV infection among PWID have largely focused on the impacts of interventions on a single disease.^7–9,11,16–28^ Broadly, these analyses find that increased levels of harm reduction strategies, testing, and treatment are required to meet goals to end the HIV epidemic or eliminate HCV infection, and that such investments are cost-effective at commonly accepted thresholds. Furthermore, combining multiple harm reduction strategies with testing and treatment of infections provides additional benefits. A smaller number of models have examined the effects of interventions on multiple infections among PWID.^29–32^ These multi-disease models align with mounting efforts to prioritize whole-person, integrated care that addresses drivers of syndemics.^33–36^

Our study adds to the existing literature by developing a novel agent-based network model of HIV and HCV transmission among PWID. The network component of the model is important to capturing heterogeneity in transmission risks, and we include interacting networks of sexual and injection equipment-sharing partnerships to reflect multiple routes of infection transmission. The model incorporates data from the National HIV Behavioral Surveillance (NHBS) system on network structure, behavioral risk factors, infection prevalence, and prevention interventions. We leveraged this model to explore strategies to achieve HIV and HCV infection incidence targets among PWID, simulating the short-term and long-term impacts of scaling single and combined interventions that have three mechanisms of action: 1) reducing the probabilities of infection transmission, 2) increasing drug injection cessation rates, and 3) treating infections.

## Methods

### Overview

We developed a stochastic agent-based network model of HIV and HCV transmission among PWID (Figure 1). This model extends an existing model that focused on HCV transmission via injection equipment sharing among PWID.^37^ Our network model includes two layers that dynamically change over time: one layer for injection equipment sharing and a second layer for sexual partnerships. We parameterized the model with data from the 2018 NHBS among PWID^38^, supplemented with estimates from the scientific literature, to represent an average urban network of 1000 PWID at simulation initiation and an additional 240 PWID entering over 10 years.

**Figure 1.**
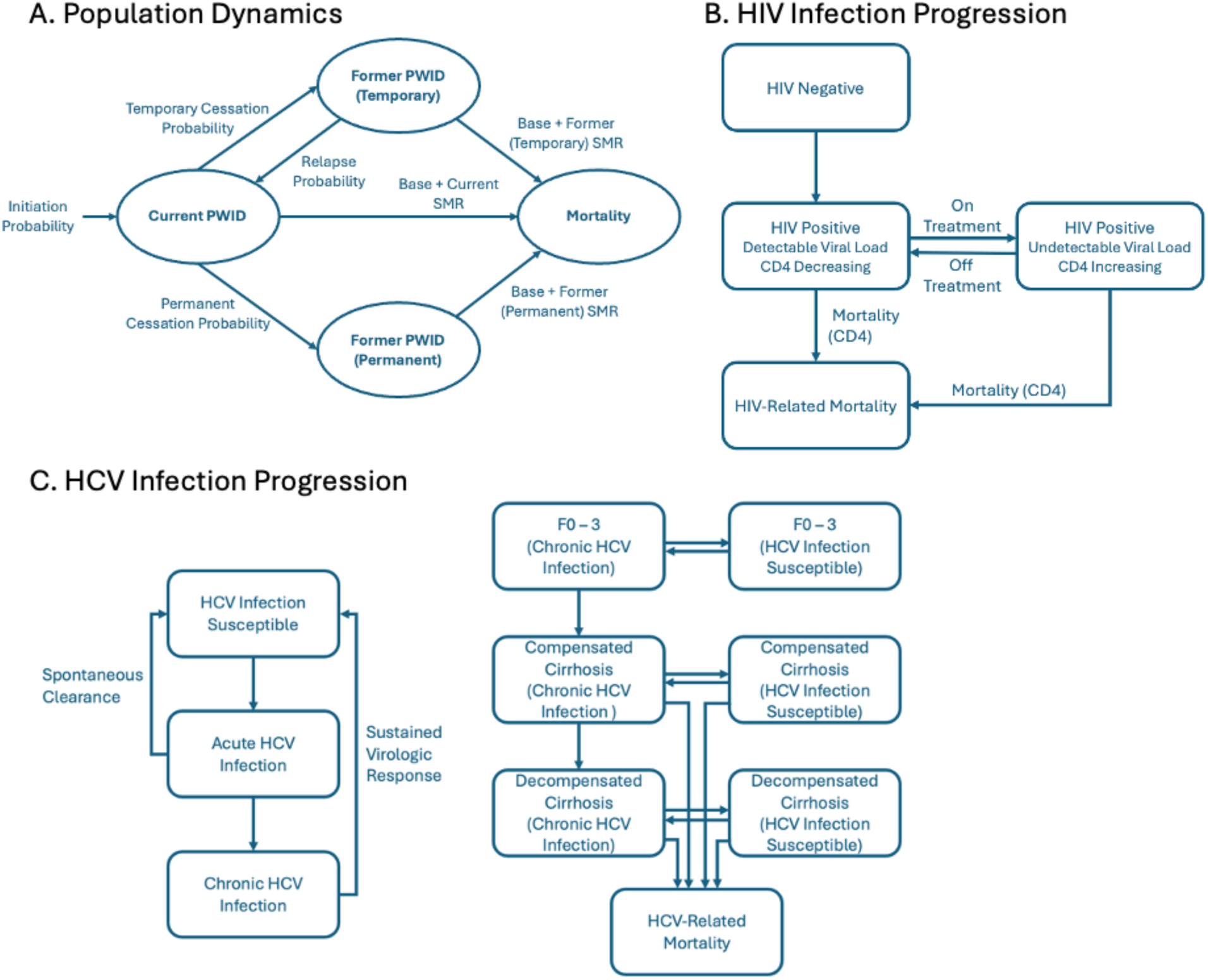
Schematic of model structure for A) population dynamics, B) HIV Infection Progression, and C) HCV Infection Progression. *Figure footnote: PWID=people who inject drugs, SMR= standardized mortality ratio, CD4= CD4+ T cell count, Mortality(CD4)=mortality as a function of a CD4+ T cell count, F0-3=METAVIR fibrosis staging scores F0 through F3. Individuals can transition from chronic HCV to HCV susceptible through treatment leading to sustained virologic response and they can transition from HCV susceptible to acute and chronic HCV infection through infection or reinfection. Fibrosis progression only occurs when an individual has a chronic HCV infection. The model captures fibrosis progression due to chronic HCV infection only (not due to other causes). Current and Former PWID statuses are based on injection drug use in the past 30 days*.

We simulated population dynamics, network partnerships, infection transmission, disease natural history, and HIV and HCV infection testing and treatment. We compared a baseline scenario calibrated to produce stable infection incidence and prevalence to alternative scenarios that decreased transmission probabilities through prevention interventions, increased drug injection cessation rates, increased HIV and HCV infection treatment rates, and decreased excess mortality rates due to injection drug use.

All analyses were conducted using R version 4.2.0 (R Project for Statistical Computing). Analytic code is available at https://github.com/PPML/multiplex. For relevant components, we followed the CHEERS reporting guideline (Appendix pp. 24-26).

### Data

Model parameter estimates and sources are reported in Table 1. Additional details on parameter estimation are described in the Appendix (pp. 3-14). When possible, we used data from the 2018 NHBS PWID cycle to represent the study population. The 2018 NHBS PWID cycle was conducted in 23 urban areas across the United States and Puerto Rico, which accounted for about two thirds of HIV diagnoses in the United States.^39^ The survey cycle included over 11,400 participants who were recruited using respondent-driven sampling (RDS).^40^ RDS is a chain recruitment method that begins with a set of seeds who recruit members of their social networks to participate in project activities, who in turn recruit other members of their social networks. Activities for NHBS were approved by the Centers for Disease Control and Prevention (NHBS was determined to be a routine disease surveillance activity) and by applicable local institutional review boards in each participating city.^39^

**Table 1.** Model parameters and sources

| Parameters | Values | References |
| --- | --- | --- |
| **Injection drug use dynamics** |  |  |
| Population size at simulation start<sup>a</sup> | 1000 | Assumed |
| Monthly number of PWID initiating and entering the simulation<sup>b</sup> | 4 | Calibrated to achieve 25% growth in the PWID population over 10 years. |
| Monthly drug injection cessation rate | 0.014 | 41 |
| Monthly drug injection relapse rate | 0.033 | 41 |
| Probability of permanent drug injection cessation<sup>c</sup> | 0.13 | 42 |
| **Mortality** |  |  |
| Standardized mortality ratio for former PWID<sup>d,e</sup> | Age-Specific | 43 |
| Standardized mortality ratio for current PWID<sup>d,e</sup> | Age-Specific | 43 |
| Additional monthly mortality rate for compensated cirrhosis (METAVIR F4) | 0.00046 | 44 |
| Additional monthly mortality rate for decompensated cirrhosis | 0.0063 | 44 |
| Post-sustained virologic response (SVR) mortality rate ratio compensated (METAVIR F4) and decompensated cirrhosis | 0.29 | 45 |
| Additional mortality rate for HIV | Function of CD4 | 46 |
| **Network simulations** |  |  |
| Baseline HCV antibody positivity prevalence | 62% | 47 |
| Baseline HCV infection prevalence | 44% | 47 |
| Baseline HIV infection prevalence | 9% | 47 |
| Baseline HIV/HCV coinfection prevalence | 4% | 47 |
| Mean number of equipment sharing partners (past 12 months) | 4.5 | 38 |
| Mean number of sexual partners (past 12 months) | 6.5 | 38 |
| Percentage of PWID with no equipment sharing partners (past 12 months) | 35% | 38 |
| Percentage of PWID with no sexual partners (past 12 months) | 15% | 38 |
| Percentage of equipment sharing partnerships that are also sexual partnerships (last equipment sharing partner) | 21% | 38 |
| Percentage of sexual partners that are “probably” or “definitely” PWID (last sexual partner) | 54% | 38 |
| Mean equipment-sharing partnership duration (years)<sup>f</sup> | 3 | Assumed |
| Mean sexual partnership duration (years) (last sexual partner) | 4 | 38 |
| Baseline prevalence of receipt of syringes from a syringe services program | 53% | 38 |
| Baseline prevalence of consistent condom use | 20% | 38 |
| **HCV transmission, treatment, and progression** |  |  |
| Monthly HCV transmission probability in discordant equipment sharing partnerships | 0.006 | Calibrated |
| Spontaneous clearance of acute HCV infection probability | 0.25 | 48–50 |
| Monthly fibrosis progression probability F0-F1 | 0.008877 | 51,52 |
| Monthly fibrosis progression probability F1-F2 | 0.00681 | 51,52 |
| Monthly fibrosis progression probability F2-F3 | 0.0097026 | 51,52 |
| Monthly fibrosis progression probability F3-F4 | 0.0096201 | 51,52 |
| Monthly fibrosis progression probability F4-decomp | 0.0097026 | 51,52 |
| Monthly HCV infection testing probability | 0.008 | 9,38,53,54 |
| HCV infection treatment probability, conditional on testing positive for HCV infection | 0.23 | 55,56 |
| HCV infection treatment completion probability, conditional on treatment initiation | 0.96 | 57–59 |
| HCV infection cure probability, conditional on treatment completion (i.e., SVR% after treatment completion) | 0.94 | 60 |
| **HIV transmission, treatment, and progression** |  |  |
| Monthly HIV transmission probability in discordant partners via equipment sharing | 0.002 | Calibrated |
| Monthly HIV transmission probability via unprotected sex with a discordant PWID partner | 0.001 | Calibrated |
| Monthly HIV transmission probability via unprotected sex with non-PWID partners<sup>g</sup> | 0.00001 | Calibrated |
| Initial CD4+ T cell count (cells/mm<sup>3</sup>) | 870 | 61–63 |
| Monthly decline in CD4+ T cell when not on treatment (cells/mm<sup>3</sup>) | 4.7 | 62–64 |
| Monthly increase in CD4+ T cell when on treatment (first six months) (cells/mm<sup>3</sup>) | 22.7 | 62,63,65 |
| Monthly increase in CD4+ T cell when on treatment (7-36 months) (cells/mm<sup>3</sup>) | 13.3 | 62,63,65 |
| Annual HIV infection testing probability | 0.55 | 38 |
| Antiretroviral therapy treatment probability among diagnosed | 0.62 | 66 |
| Annual probability of stopping antiretroviral therapy | 0.30 | 67 |
| **Health utility/quality of Life (QoL)** |  |  |
| Base (age 14 – 29 y) | 0.928 | 68 |
| Base (age 30 – 39y) | 0.918 | 68 |
| Base (age 40 – 49y) | 0.887 | 68 |
| Base (age 50 – 59y) | 0.861 | 68 |
| Base (age 60 – 69y) | 0.840 | 68 |
| Base (age 70 – 79y) | 0.802 | 68 |
| Base (age 80 – 100y) | 0.782 | 68 |
| Current PWID | 0.68 | 69 |
| Former PWID | 0.82 | 69 |
| METAVIR F0-F3, with chronic HCV infection | 0.96 | 45,70 |
| Compensated cirrhosis, with chronic HCV infection | 0.85 | 45,70 |
| Decompensated cirrhosis, with chronic HCV infection | 0.77 | 45,70 |
| METAVIR F0-F3 at time of treatment, with SVR | 1 | 45,70 |
| Compensated cirrhosis at time of treatment, with SVR | 0.96 | 45,70 |
| Decompensated cirrhosis at time of treatment, with SVR | 0.93 | 45,70 |
| CD4+ T cell count ≥ 500 cells/mm<sup>3</sup> | 1 | 71–73 |
| CD4+ T cell count ≥ 200 cells/mm<sup>3</sup> & CD4+ T cell count < 500 cells/mm<sup>3</sup>, on antiretroviral therapy and virally suppressed | 0.980 | 71–73 |
| CD4+ T cell count ≥ 200 cells/mm<sup>3</sup> & CD4+ T cell count < 500 cells/mm<sup>3</sup>, not on antiretroviral therapy | 0.867 | 71–73 |
| CD4+ T cell count < 200 cells/mm<sup>3</sup>, on antiretroviral therapy and virally suppressed | 0.799 | 71–73 |
| CD4+ T cell count < 200 cells/mm<sup>3</sup>, not on antiretroviral therapy | 0.707 | 71-73 |

<sup>a</sup> Prior analysis suggested that different network sizes (n=500 and n=2000, versus n=1000) had minimal effects on results. Our assumed network size is included as a limitation, and we varied the number of PWID initiating injection drug use in sensitivity analyses.<sup>37</sup>
<sup>b</sup> Assumed a slowing in the rate of growth of the PWID population compared to 2010-2019 (Appendix pp. 3).
<sup>c</sup> Conditional probability among those stopping injection drug use in a given month.
<sup>d</sup> Current and Former PWID statuses are based on injection drug use in the past 30 days.
<sup>e</sup> Details on the age-specific estimates of excess mortality due to current and former injection drug use are included in the Appendix (pp. 6-7).
<sup>f</sup> Among equipment sharing partnerships in two studies, the average duration of acquaintance between partners was 10 years.
<sup>g</sup> Assumed homogenous mixing with all non-PWID adults. Adjusted the monthly probability of transmission via unprotected sex with a discordant PWID partner for the average number of non-PWID sexual partners, the prevalence of HIV infection among adults in the United States, and the probability of viral suppression among people with HIV.

### Network Simulation

We used separable temporal exponential-family random graph models (STERGM) to simulate networks of injection equipment sharing and sexual partnerships that matched observed network summary statistics from 2018 NHBS data for PWID (Table 1), which was the most recent NHBS PWID data available at the time of the analysis. These dynamic network models allow for partnership formation and dissolution over time. We used the Statnet suite of R packages for estimation of STERGM coefficients and network simulation.^74^ We fit models to produce networks of sexual and injection equipment-sharing partnerships that matched the mean number of partners and percentage of PWID with zero partners. Additionally, we informed the overlap of the equipment sharing and sexual networks based on the percentage of last equipment sharing partners who were also sexual partners, and the percentage of last sexual partners who were “probably or definitely PWID.” Details on the construction and assumptions underlying these network targets are described in the Appendix (pp. 14-16).

### Infection Transmission, Natural History, and Treatment

We modeled transmission of HCV through injection equipment sharing, and transmission of HIV through injection equipment sharing and sexual activity. We calibrated monthly probabilities of transmission to HIV and HCV infection prevalence from NHBS.^47^ To calibrate these unknown parameters, we implemented a grid search to identify the parameter set that minimized the mean absolute percentage error between model estimates and calibration targets (Appendix pp. 16-18). We modeled the natural history of HIV and HCV infection, as well as the impact of treatment on individuals’ health and risk of transmission. In our main analysis, we assumed that individuals on antiretroviral therapy for HIV who reach viral suppression have no potential to transmit HIV via either injection equipment sharing or sexual activity.^75^

### Alternative Scenarios

We explored the impacts of three interventions at three levels of coverage (baseline, moderate, and high). The first, referred to as “Prevention Interventions,” reduced the probability of infection transmission. Individuals covered with prevention interventions had 50% lower probabilities of HIV and HCV transmission through injection equipment sharing, and 80% lower probabilities of HIV transmission through sex. These reductions were based on estimates of the effectiveness of programs that provide safer injection equipment and the effectiveness of consistent use of male condoms.^76,77^ For transmission through injection equipment sharing, the baseline level of coverage (53%) was chosen to match the percentage of PWID in NHBS receiving syringes from syringe services programs in the past 12 months. For transmission through sex, the baseline level of coverage (20%) was the approximate percentage of PWID in NHBS reporting consistently using condoms. We defined moderate coverage of the prevention interventions as a 15 percentage-point increase from baseline, and high coverage as a 30 percentage-point increase.

The second intervention, “Cessation,” increased the baseline drug injection cessation rate of 16 per 100 person-years, to 31 per 100 person-years at moderate levels, and 46 per 100 person-years at high levels. The rate of relapse and the probability of permanent drug injection cessation among those ceasing injection drug use in a given month remained unchanged across scenarios. This intervention illustrated some of the effects of increasing access to medications for opioid use disorder (MOUD).

The third intervention, “Test and Treat,” increased the probability of testing and treating PWID with current HCV infection, and increased the duration of viral suppression (and, as a result, the proportion suppressed at any time point) among individuals with HIV. We assumed HIV testing probabilities remained constant at the baseline level in all scenarios. For HCV, the monthly probability of testing was 0.008 at the baseline intervention level, 0.07 at the moderate intervention level, and 0.18 at the high intervention level. The probability of treatment initiation after diagnosis was 0.23 at the baseline intervention level, 0.38 at the moderate intervention level, and 0.53 at the high intervention level. This resulted in an estimated 3% of people with current HCV infection achieving sustained virologic response (SVR) per year in the baseline, while 16% and 28% achieved SVR yearly at moderate and high intervention levels, respectively. For HIV, the annual probability of stopping antiretroviral therapy was decreased from 0.3 in the baseline, to 0.14 at moderate intervention coverage, and to 0.02 at high coverage. In the baseline, an estimated 44% of all individuals with HIV were virally suppressed during the 10-year intervention period, compared to 58% at the moderate intervention level and 71% at the high intervention level.

We also explored the effects of reducing non-hepatitis C and non-HIV excess mortality due to drug use by 25% or 50%, which is illustrative of some of the effects of increasing access to overdose prevention including overdose education, naloxone distribution, and MOUD. Drug-related deaths, an important competing risk for mortality among PWID, could modulate the realized health benefits of achieving elimination targets.

### Analytic Outcomes

Our primary outcome measures were the percentage reductions in new HIV and HCV infections (including HCV reinfections), compared to baseline, during the 10-year intervention period (2019-2028). These outcome measures approximated the incidence targets included in national HIV and HCV strategic plans.^1,2^ Additionally, we quantified the long-term health impacts of different strategies for elimination using life-years and quality-adjusted life-years calculated over an 80-year follow-up period, with or without discounting at 3% per year. We used a consistent population size (n=1240) to report per-person life-years and quality-adjusted life-years, which weighted the simulated PWID population by their potential intervention exposure duration over the 10-year intervention period (Appendix pp. 5).

We recorded mortality attributable to four categories of causes of death: drug-related, hepatitis C-related, HIV-related, and deaths due to other causes, referred to as background deaths. Drug-related deaths included fatal overdoses, as well as other contributors to excess mortality among PWID (Appendix pp. 6-8).

To compute quality-adjusted life-years, we estimated health utilities over each of five attributes, including age group, injection drug use status (current vs. former), HCV infection status (SVR vs. no SVR), liver fibrosis stage (METAVIR stages F0-F3, compensated cirrhosis, decompensated cirrhosis), and HIV infection status (by CD4 cell count category and antiretroviral therapy treatment status). We assumed health utilities were independent across the five attributes, which resulted in a multiplicative model for overall utility, and we adjusted published estimates to avoid double-counting utility decrements (Appendix pp. 12-14).

We captured stochastic uncertainty across 150 simulation repetitions. We used common random numbers to reduce the impact of stochastic noise on scenario comparisons (Appendix pp. 18-20).^78^ Our 95% uncertainty intervals (UI) are based on the 2.5^th^ and 97.5^th^ percentile of simulation repetitions.

### Sensitivity Analyses

We ran targeted one-way sensitivity analyses for uncertain parameters to examine the stability of results. We included sensitivity analyses for HIV and HCV transmission probabilities, the proportion of the population that can be reached by testing, the effectiveness of antiretroviral therapy to prevent HIV transmission through injection equipment sharing, the reduction in risk of reinfection following hepatitis C treatment,^79–82^ the probability of HCV infection treatment completion,^83^ and the number of individuals initiating injection drug use each month.

## Results

In the baseline scenario, we estimated 448 (95% UI: 404-496) new HCV infections and 50 (95% UI: 35-70) new HIV infections over ten years. Simulated prevalence of HCV infection was 44% (95% UI: 41-47%), prevalence of HIV infection was 8% (95% UI: 7-10%), and prevalence of HIV/HCV co-infection was 5% (95% UI: 4%-6%). Over an 80-year follow-up period, PWID in the analytic cohort accrued 20.2 (95% UI: 19.7-20.5) discounted life-years, 36.0 (95% UI: 34.6-37.0) undiscounted life-years, 12.1 (95% UI: 11.8-12.3) discounted quality-adjusted life-years, and 21.5 (95% UI: 20.7-22.3) undiscounted quality-adjusted life-years. In the baseline scenario, the mean age at death was 66.5 (95% UI: 65.5-67.6) years, compared to a life expectancy at birth of 77.5 for the general US population.^84^ Forty-five percent (95% UI: 42-48%) of deaths in the baseline scenario were drug-related deaths.

Expanding coverage of the three interventions resulted in substantial reductions in both HIV and HCV incidence among PWID over a ten-year time horizon (Figure 2). At the levels of intervention coverage examined in this analysis, combinations of interventions were required to reduce new HIV and new HCV infections by 90%. High coverage across all three intervention strategies yielded an 86% (95% UI: 72-96%) decrease in new HIV infections and a 90% (95% UI: 87-94%) decrease in new HCV infections over ten years among PWID. At high levels of coverage, deaths due to HIV decreased by 27% (95% UI: 9-46%), and deaths due to HCV infection decreased by 72% (95% UI: 65-81%). Moderate coverage across all three strategies yielded 62% (95% UI: 39-81%) and 68% (95% UI: 61-74%) decreases in new HIV and HCV infections among PWID, respectively.

**Figure 2.**
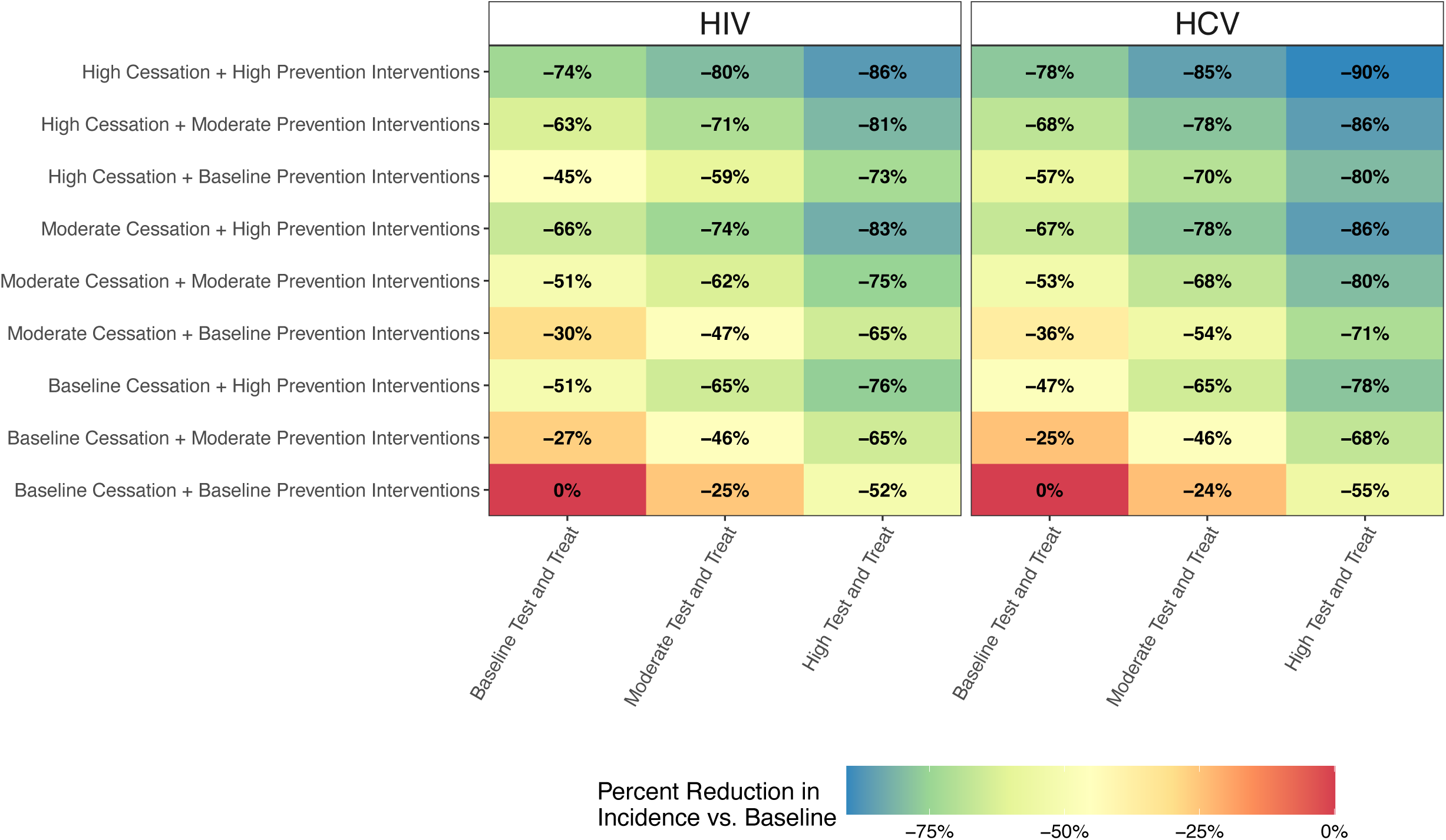
Impacts of scaling single and combined interventions on HIV and HCV infection incidence among PWID, compared to baseline. Footnote: Baseline cessation rate is 16 per 100 person-years, moderate cessation rate is 31 per 100 person-years, and high cessation rate is 46 per 100 person-years. Baseline prevention intervention coverage for injection equipment sharing is 53%, moderate coverage is 68%, and high coverage is 83%. Baseline prevention intervention coverage for sex is 20%, moderate coverage is 35%, and high coverage is 50%. The percentage of people with HCV infection reaching SVR per year is 3% at baseline test and treat, 16% at moderate test and treat, and 28% at high test and treat. The percentage of people with HIV that are virally suppressed is 44% at baseline test and treat, 58% at moderate test and treat, and 71% at high test and treat.

At these moderate and high levels of intervention coverage, combinations of interventions were largely additive or sub-additive in their effects on incidence and prevalence. At lower increases in intervention coverage, combining testing and treatment with prevention and cessation interventions in some cases produced super-additive, or synergistic, effects (Appendix pp. 21, Supplemental Figure S14).

Drug-related deaths accounted for the largest proportion of cause-specific deaths among PWID. Increasing cessation emerged as the most effective single intervention to increase quality-adjusted life-years among PWID (Figure 3). Increasing cessation reduces drug-related deaths and reduces transmission of both HIV and HCV infection. Reaching moderate levels of cessation resulted in an increase of 0.8 (95% UI: 0.6-0.9) discounted quality-adjusted life-years, compared to baseline levels. This increase was larger than that achieved by combining high test and treat with high prevention intervention coverage (0.7, 95% UI: 0.6-0.8). Moderate levels across all three interventions yielded an increase of 1.2 (95% UI: 1.1-1.4) discounted quality-adjusted life-years.

**Figure 3.**
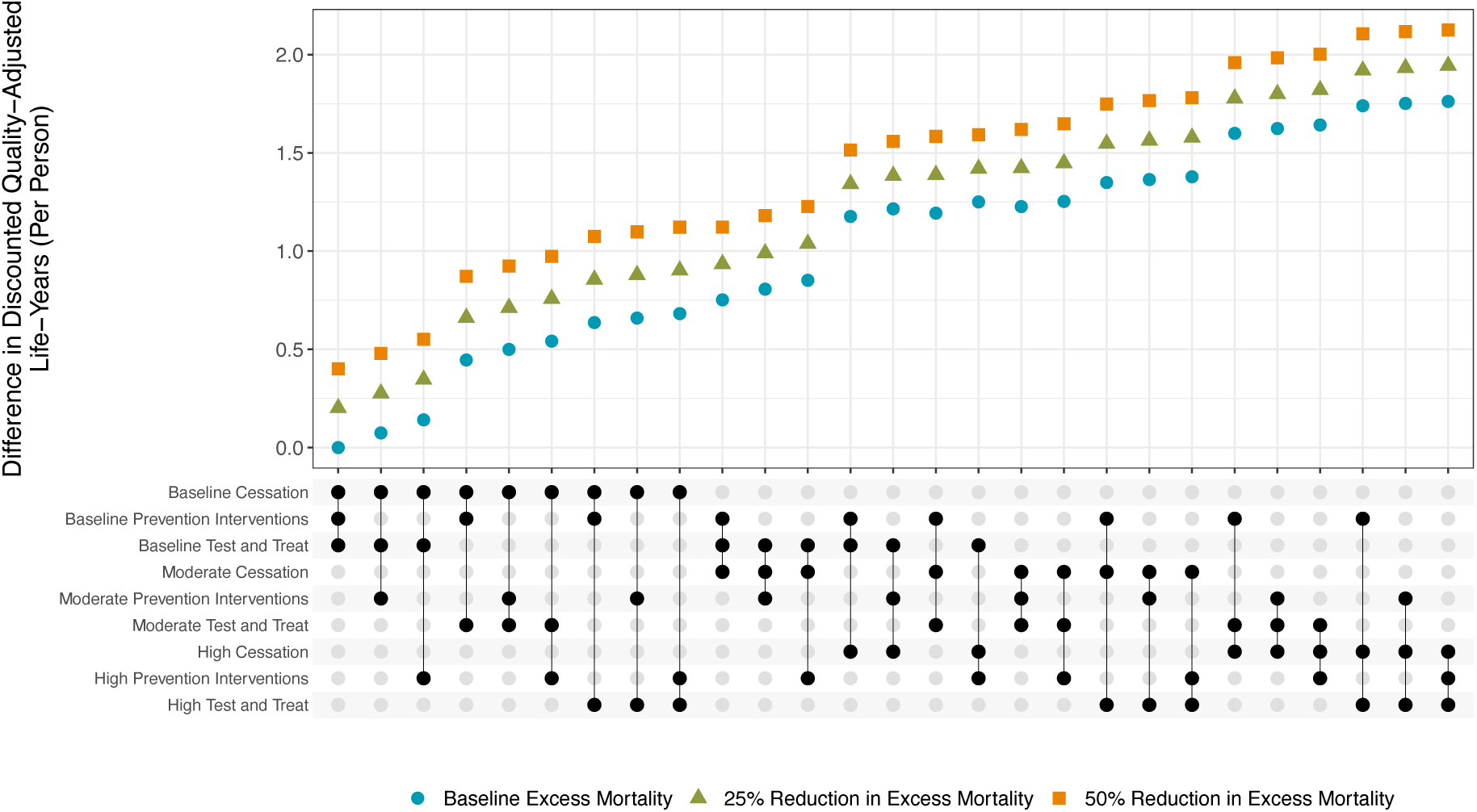
Difference in discounted quality-adjusted life-years per person, compared to baseline. Footnote: Per-person estimates are based on a consistent population size of 1240. Estimates are discounted at a rate of 3% per year. Baseline cessation rate is 16 per 100 person-years, moderate cessation rate is 31 per 100 person-years, and high cessation rate is 46 per 100 person-years. Baseline prevention intervention coverage for injection equipment sharing is 53%, moderate coverage is 68%, and high coverage is 83%. Baseline prevention intervention coverage for sex is 20%, moderate coverage is 35%, and high coverage is 50%. The percentage of people with HCV infection reaching SVR per year is 3% at baseline test and treat, 16% at moderate test and treat, and 28% at high test and treat. The percentage of people with HIV that are virally suppressed is 44% at baseline test and treat, 58% at moderate test and treat, and 71% at high test and treat. Excess mortality refers to excess mortality due to current or former injection drug use.

Across sensitivity analyses, variation in parameter values impacted the estimated baseline HIV and hepatitis C incidence rates (Figure 4, top). A lower effectiveness of HIV treatment in reducing transmission through injection equipment sharing and a lower HCV infection treatment completion probability decreased the incremental benefits of scaling testing and treatment on HIV and hepatitis C incidence, respectively (Figure 4, bottom). Expanding the proportion of the population that can be reached by testing resulted in greater reductions in HIV and hepatitis C incidence, particularly at the high intervention level, whereas reducing the proportion of the population that can be reached by testing resulted in smaller reductions in incidence. Including a reduction in risk of reinfection after treatment of HCV infection resulted in greater benefits from treatment on hepatitis C incidence. Drug injection initiation rates predominantly altered the PWID population size and impacted infection incidence by changing the number of susceptible individuals entering the modeled population. The effects of the prevention and cessation intervention strategies were least sensitive to the parameters included in sensitivity analyses.

**Figure 4.**
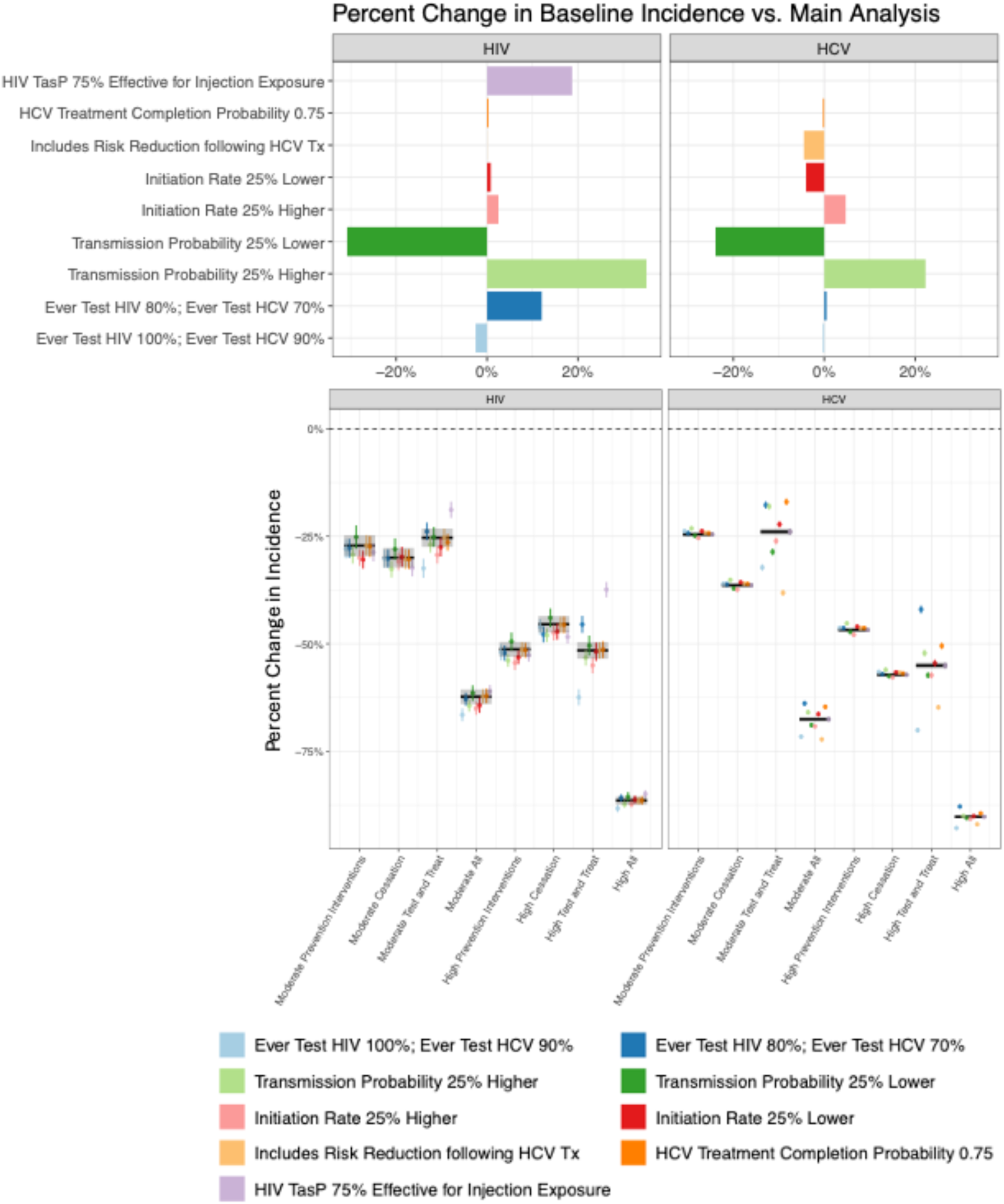
Variation in estimated relative differences in HIV and HCV infection incidence among PWID across sensitivity analyses. Footnote: Baseline cessation rate is 16 per 100 person-years, moderate cessation rate is 31 per 100 person-years, and high cessation rate is 46 per 100 person-years. Baseline prevention intervention coverage for injection equipment sharing is 53%, moderate coverage is 68%, and high coverage is 83%. Baseline prevention intervention coverage for sex is 20%, moderate coverage is 35%, and high coverage is 50%. The percentage of people with HCV infection reaching SVR per year is 3% at baseline test and treat, 16% at moderate test and treat, and 28% at high test and treat. The percentage of people with HIV that are virally suppressed is 44% at baseline test and treat, 58% at moderate test and treat, and 71% at high test and treat. TasP refers to the reduction in HIV transmission for injection equipment sharing due to viral suppression from antiretroviral therapy. HCV Tx refers to treatment of HCV infection with direct-acting antiviral medication. In the bottom figure, the black line is the main analysis estimate. Uncertainty displayed is the standard error of the mean estimate.

## Discussion

Substantial improvements in survival and health-related quality of life for PWID can be achieved by scaling combinations of intervention strategies. Using a stochastic, agent-based network model of HIV and HCV transmission among PWID, we found that a combined strategy that sustained coverage of prevention, cessation, and test and treat interventions at high levels for 10 years could potentially decrease the incidence of HIV by 86% and HCV infection by 90% and produce an increase of 1.8 discounted quality-adjusted life-years per person. Scaled to an estimated total population of 3.7 million PWID in the United States,^5^ sustained high coverage across the three interventions could avert 1.33 million HCV infections and 0.15 million HIV infections over 10 years. Among the three strategies examined in our study, increasing cessation of injection drug use was the most effective single mechanism, when considering effects on both infection incidence and quality-adjusted life-years. Our analysis complements existing literature by reinforcing the value of comprehensive services for PWID using a model parameterized with 2018 NHBS data from 23 US cities, that captures impacts across multiple infections, and that incorporates heterogeneity in risk of infection due to network structure.

States and local jurisdictions have an opportunity to use opioid settlement funding to turn the tide on the ongoing syndemic of substance use, hepatitis C, HIV, and other STIs.^85^ Our model-based analysis suggests that investments to expand access to harm reduction services could yield large benefits across multiple outcomes. For example, increasing access to and use of MOUD can reduce the rate of overdose, reduce transmission of infections, and improve health-related quality of life for PWID. The top six combinations of intervention strategies that generated the highest per-person QALY gains included high levels of cessation. While outside the scope of this analysis, multi-disease syndemic models could be used to quantify the wide-ranging benefits of investments targeting social determinants of health, such as housing, that act further upstream.^86,87^ Capturing the totality of downstream impacts is critical to appropriately valuing investments in upstream interventions.^88^

The results of our analyses should be interpreted in the context of multiple limitations. First, we did not model specific interventions. Our analysis focused on mechanisms through which multiple candidate interventions could act, prompting further work to model the effects and cost-effectiveness of specific, evidence-based interventions. Second, our modeled network of PWID was smaller than the estimated size of the PWID population in many urban areas.^5,89,90^ While prior studies have found that the effects of interventions are not sensitive to network size, additional unobserved features of network structure, such as the number of components and their connectivity, may be important.^37^ In addition, the resources required to expand coverage of interventions may not scale linearly with network size. Third, we parameterized our model to reflect an average NHBS project area. NHBS project areas are heterogeneous in terms of their demographics, prevalence of infections, and access to harm reduction services.^39,47^ Our study provides a foundation for future work to incorporate local data that may be more informative to policymakers at the local level.^6^ Additionally, data from NHBS, in which participants are recruited through RDS, are not nationally representative and might not be generalizable to all US urban areas, nonurban areas, or all PWID.^91^ However, the hidden and hard-to-reach nature of this population prevents collection of nationally representative samples. NHBS data represent the gold standard of national level data used to inform HIV prevention among this population in the United States.^38^ Further, we used data collected prior to the COVID-19 pandemic and did not seek to incorporate its effects. Fourth, measures to define network overlap and partnership duration were derived from questions about individuals’ last sexual and last equipment sharing partners. These measures may not be representative of all such partners. Additionally, we did not incorporate correlations between partnership duration and number of partners for either sexual or equipment sharing partnerships. Capturing additional heterogeneity in partnership types will improve the fidelity of our simulated network to true networks among PWID. Fifth, we did not include heterogeneity in behaviors or coverage of interventions by sex, race/ethnicity, or other sociodemographic characteristics, and did not examine distributional impacts of interventions. Sixth, we did not quantify benefits from reduced HIV transmission between PWID and their non-PWID sexual partners. Seventh, we did not specifically model drug use through routes other than injection. Although the transition from injection to other routes of drug use can decrease infectious disease transmission, substantial health risks including overdose can persist.^92,93^ Finally, due to the computational intensity of agent-based network models, we did not attempt to run full probabilistic sensitivity analyses or explore the implications of joint uncertainty in model parameters. Comparative modeling using other approaches, such as compartmental or microsimulation models, would permit further exploration of these limitations.^94^

Our study underscores the potential value of implementing and expanding evidence-based comprehensive treatment and harm reduction services for PWID. The benefits span decreased HIV and HCV infection incidence and prevalence, and increased survival and quality of life among PWID. We found that increasing drug injection cessation, which can be achieved by expanding access to and uptake of MOUD, is the most effective single intervention. We also found that combinations of interventions, implemented to achieve ambitious levels of coverage, were required to decrease HIV and HCV infection incidence by 90%. Considering that the feasibility of scaling interventions is likely non-linear, with increasing resource requirements to reach very high levels of coverage, our analysis suggests that investing in a broad package of interventions may be more fruitful than myopically investing in any single intervention.

## Data Availability

Analytic code is available at https://github.com/PPML/multiplex. National HIV Behavioral Surveillance data are not publicly available.

## Acknowledgments

**NHBS Study Group: Atlanta, GA:** Pascale Wortley, Jeff Todd, David Melton; **Baltimore, MD:** Colin Flynn, Danielle German; **Boston, MA:** Monina Klevens, Rose Doherty, Conall O’Cleirigh; **Chicago, IL:** Antonio D. Jimenez, Thomas Clyde; **Dallas, TX:** Jonathon Poe, Margaret Vaaler, Jie Deng; **Denver, CO:** Alia Al-Tayyib, Daniel Shodell; **Detroit, MI:** Emily Higgins, Vivian Griffin, Corrinne Sanger; **Houston, TX:** Salma Khuwaja, Zaida Lopez, Paige Padgett; **Los Angeles, CA:** Ekow Kwa Sey, Yingbo Ma, Hugo Santacruz; **Memphis, TN:** Meredith Brantley, Christopher Mathews, Jack Marr; **Miami, FL:** Emma Spencer, Willie Nixon, David Forrest; **Nassau-Suffolk, NY:** Bridget Anderson, Ashley Tate, Meaghan Abrego; **New Orleans, LA:** William T. Robinson, Narquis Barak, Jeremy M. Beckford; **New York City, NY:** Sarah Braunstein, Alexis Rivera, Sidney Carrillo **Newark, NJ:** Abdel R. Ibrahim, Afework Wogayehu, Luis Moraga; **Philadelphia, PA:** Kathleen A. Brady, Jennifer Shinefeld, Chrysanthus Nnumolu,; **Portland, OR:** Timothy W. Menza, E. Roberto Orellana, Amisha Bhattari; **San Diego, CA:** Anna Flynn, Onika Chambers, Marisa Ramos; **San Francisco, CA:** Willi McFarland, Jessica Lin, Desmond Miller; **San Juan, PR:** Sandra Miranda De León, Yadira Rolón-Colón, María Pabón Martínez; **Seattle, WA:** Tom Jaenicke, Sara Glick; **Virginia Beach, VA:** Jennifer Kienzle, Brandie Smith, Toyah Reid; **Washington, DC:** Jenevieve Opoku, Irene Kuo; **CDC:** Monica Adams, Christine Agnew Brune, Amy Baugher, Dita Broz, Janet Burnett, Susan Cha, Johanna Chapin-Bardales, Paul Denning, Teresa Finlayson, Senad Handanagic, Terence Hickey, Dafna Kanny, Kathryn Lee, Rashunda Lewis, Elana Morris, Evelyn Olansky, Taylor Robbins, Catlainn Sionean, Amanda Smith, Anna Teplinskaya, Lindsay Trujillo, Cyprian Wejnert, Ari Whiteman, Mingjing Xia.

## References

1. U.S. Department of Health and Human Services. Viral Hepatitis National Strategic Plan for the United States: A Roadmap to Elimination (2021-2025), https://www.hhs.gov/sites/default/files/Viral-Hepatitis-National-Strategic-Plan-2021-2025.pdf (2020).

2. The White House. National HIV/AIDS Strategy for the United States 2022–2025, https://www.whitehouse.gov/wp-content/uploads/2021/11/National-HIV-AIDS-Strategy.pdf (2021).

3. HIV Prevention in the United States: Mobilizing to End the Epidemic, https://www.cdc.gov/hiv/pdf/policies/cdc-hiv-prevention-bluebook.pdf.

4. CDC. NCHHSTP Syndemic Approach. *National Center for HIV, Viral Hepatitis, STD, and Tuberculosis Prevention*, https://www.cdc.gov/nchhstp/about/syndemic.html (2024, accessed 26 September 2024).

5. Bradley H, Hall EW, Asher A, et al. Estimated Number of People Who Inject Drugs in the United States. Clin Infect Dis Off Publ Infect Dis Soc Am 2023; 76: 96–102.

6. Schackman BR, Meisel ZF. Commentary on Barbosa, et al. (2019): The value of using community simulation modeling to achieve HCV elimination targets in people who inject drugs. Addict Abingdon Engl 2019; 114: 2279–2280.

7. Krebs E, Zang X, Enns B, et al. Ending the HIV Epidemic Among Persons Who Inject Drugs: A Cost-Effectiveness Analysis in Six US Cities. J Infect Dis 2020; 222: S301–S311.

8. Barbosa C, Fraser H, Hoerger TJ, et al. Cost-effectiveness of scaling-up HCV prevention and treatment in the United States for people who inject drugs. Addict Abingdon Engl 2019; 114: 2267–2278.

9. Blake A, Smith JE. Modeling Hepatitis C Elimination Among People Who Inject Drugs in New Hampshire. JAMA Netw Open 2021; 4: e2119092.

10. Boodram B, Mackesy-Amiti ME, Khanna A, et al. People who inject drugs in metropolitan Chicago: A meta-analysis of data from 1997-2017 to inform interventions and computational modeling toward hepatitis C microelimination. PloS One 2022; 17: e0248850.

11. Bellerose M, Zhu L, Hagan LM, et al. A review of network simulation models of hepatitis C virus and HIV among people who inject drugs. Int J Drug Policy 2021; 88: 102580.

12. Cassels S, Clark SJ, Morris M. Mathematical Models for HIV Transmission Dynamics. J Acquir Immune Defic Syndr 1999 2008; 47: S34–S39.

13. Monteiro JFG, Escudero DJ, Weinreb C, et al. Understanding the effects of different HIV transmission models in individual-based microsimulation of HIV epidemic dynamics in people who inject drugs. Epidemiol Infect 2016; 144: 1683–1700.

14. Cipriano LE, Goldhaber-Fiebert JD. Population Health and Cost-Effectiveness Implications of a ‘Treat All’ Recommendation for HCV: A Review of the Model-Based Evidence. MDM Policy Pract 2018; 3: 2381468318776634.

15. Weinstein MC, O’Brien B, Hornberger J, et al. Principles of good practice for decision analytic modeling in health-care evaluation: report of the ISPOR Task Force on Good Research Practices--Modeling Studies. Value Health J Int Soc Pharmacoeconomics Outcomes Res 2003; 6: 9–17.

16. Siegele-Brown C, Siegele-Brown M, Cook C, et al. Testing to sustain hepatitis C elimination targets in people who inject drugs: A network-based model. J Viral Hepat 2023; 30: 242– 249.

17. Tatara E, Gutfraind A, Collier NT, et al. Modeling hepatitis C micro-elimination among people who inject drugs with direct-acting antivirals in metropolitan Chicago. PloS One 2022; 17: e0264983.

18. Brown C, Siegele M, Wright M, et al. Injecting network structure determines the most efficient strategy to achieve Hepatitis C elimination in people who inject drugs. J Viral Hepat 2021; 28: 1274–1283.

19. Stocks T, Martin LJ, Kühlmann-Berenzon S, et al. Dynamic modeling of hepatitis C transmission among people who inject drugs. Epidemics 2020; 30: 100378.

20. Heffernan A, Cooke GS, Nayagam S, et al. Scaling up prevention and treatment towards the elimination of hepatitis C: a global mathematical model. Lancet Lond Engl 2019; 393: 1319–1329.

21. Scott N, Sacks-Davis R, Pedrana A, et al. Eliminating hepatitis C: The importance of frequent testing of people who inject drugs in high-prevalence settings. J Viral Hepat 2018; 25: 1472–1480.

22. Gountas I, Sypsa V, Blach S, et al. HCV elimination among people who inject drugs. Modelling pre- and post–WHO elimination era. PLOS ONE 2018; 13: e0202109.

23. Jarlais DD, Bobashev G, Feelemyer J, et al. Modeling HIV transmission among persons who inject drugs (PWID) at the ‘End of the HIV Epidemic’ and during the COVID-19 pandemic. Drug Alcohol Depend 2022; 238: 109573.

24. Zang X, Goedel WC, Bessey SE, et al. The impact of syringe services program closure on the risk of rebound HIV outbreaks among people who inject drugs: a modeling study. AIDS Lond Engl 2022; 36: 881–888.

25. Bernard CL, Owens DK, Goldhaber-Fiebert JD, et al. Estimation of the cost-effectiveness of HIV prevention portfolios for people who inject drugs in the United States: A model-based analysis. PLoS Med 2017; 14: e1002312.

26. Song DL, Altice FL, Copenhaver MM, et al. Cost-effectiveness analysis of brief and expanded evidence-based risk reduction interventions for HIV-infected people who inject drugs in the United States. PloS One 2015; 10: e0116694.

27. Fu R, Owens DK, Brandeau ML. Cost-effectiveness of alternative strategies for provision of HIV preexposure prophylaxis for people who inject drugs. AIDS Lond Engl 2018; 32: 663– 672.

28. Martin NK, Hickman M, Hutchinson SJ, et al. Combination Interventions to Prevent HCV Transmission Among People Who Inject Drugs: Modeling the Impact of Antiviral Treatment, Needle and Syringe Programs, and Opiate Substitution Therapy. Clin Infect Dis Off Publ Infect Dis Soc Am 2013; 57: S39–S45.

29. Chiosi JJ, Mueller PP, Chhatwal J, et al. A multimorbidity model for estimating health outcomes from the syndemic of injection drug use and associated infections in the United States. BMC Health Serv Res 2023; 23: 760.

30. Cheema JS, Mathews WC, Wynn A, et al. Hepatitis C Virus Micro-elimination Among People With HIV in San Diego: Are We on Track? Open Forum Infect Dis 2023; 10: ofad153.

31. Fu R, Gutfraind A, Brandeau ML. Modeling a dynamic bi-layer contact network of injection drug users and the spread of blood-borne infections. Math Biosci 2016; 273: 102–113.

32. Fraser H, Mukandavire C, Martin NK, et al. HIV treatment as prevention among people who inject drugs - a re-evaluation of the evidence. Int J Epidemiol 2017; 46: 466–478.

33. Long K, Mette E, Manz J. Tackling the Trifecta: State Approaches to Addressing Co-Occurring Substance Use Disorders, HIV, and Hepatitis C. *NASHP*, https://nashp.org/tackling-the-trifecta-state-approaches-to-addressing-co-occurring-substance-use-disorders-hiv-and-hepatitis-c/ (2020, accessed 8 June 2024).

34. Martel-Laferrière V, Feaster DJ, Metsch LR, et al. M(2)HepPrEP: study protocol for a multi-site multi-setting randomized controlled trial of integrated HIV prevention and HCV care for PWID. Trials 2022; 23: 341.

35. Bromberg DJ, Mayer KH, Altice FL. Identifying and Managing Infectious Disease Syndemics in Patients with HIV. Curr Opin HIV AIDS 2020; 15: 232–242.

36. Fukuda HD, Randall LM, Meehan T, et al. Leveraging Health Department Capacities, Partnerships, and Health Insurance for Infectious Disease Response in Massachusetts, 2014-2018. Public Health Reports® 2020; 135: 75S–81S.

37. Zhu L, Thompson WW, Hagan L, et al. Potential impact of curative and preventive interventions toward hepatitis C elimination in people who inject drugs–A network modeling study. Int J Drug Policy 2024; 130: 104539.

38. Kanny D, Broz D, Finlayson T, et al. A Key Comprehensive System for Biobehavioral Surveillance of Populations Disproportionately Affected by HIV (National HIV Behavioral Surveillance): Cross-sectional Survey Study. JMIR Public Health Surveill 2022; 8: e39053.

39. Centers for Disease Control and Prevention. HIV Infection Risk, Prevention, and Testing Behaviors among Persons Who Inject Drugs—National HIV Behavioral Surveillance: Injection Drug Use, 23 U.S. Cities, 2018. HIV Surveillance Special Report 24, https://stacks.cdc.gov/view/cdc/106349 (February 2020).

40. Heckathorn DD. Respondent-Driven Sampling II: Deriving Valid Population Estimates from Chain-Referral Samples of Hidden Populations. Soc Probl 2002; 49: 11–34.

41. Galai N, Safaeian M, Vlahov D, et al. Longitudinal patterns of drug injection behavior in the ALIVE Study cohort,1988-2000: description and determinants. Am J Epidemiol 2003; 158: 695–704.

42. Shah NG, Galai N, Celentano DD, et al. Longitudinal predictors of injection cessation and subsequent relapse among a cohort of injection drug users in Baltimore, MD, 1988-2000. Drug Alcohol Depend 2006; 83: 147–156.

43. Evans E, Li L, Min J, et al. Mortality among individuals accessing pharmacological treatment for opioid dependence in California, 2006-10. Addict Abingdon Engl 2015; 110: 996–1005.

44. Wang PL, Djerboua M, Flemming JA. Cause-specific mortality among patients with cirrhosis in a population-based cohort study in Ontario (2000–2017). Hepatol Commun 2023; 7: e00194.

45. van der Meer AJ, Veldt BJ, Feld JJ, et al. Association between sustained virological response and all-cause mortality among patients with chronic hepatitis C and advanced hepatic fibrosis. JAMA 2012; 308: 2584–2593.

46. Glaubius R, Kothegal N, Birhanu S, et al. Disease progression and mortality with untreated HIV infection: evidence synthesis of HIV seroconverter cohorts, antiretroviral treatment clinical cohorts and population-based survey data. J Int AIDS Soc 2021; 24: e25784.

47. Chapin-Bardales J, Asher A, Broz D, et al. Hepatitis C virus infection and co-infection with HIV among persons who inject drugs in 10 U.S. cities—National HIV Behavioral Surveillance, 2018. Int J Drug Policy 2024; 104387.

48. Grebely J, Raffa JD, Lai C, et al. Factors associated with spontaneous clearance of hepatitis C virus among illicit drug users. Can J Gastroenterol 2007; 21: 447–451.

49. Thomas DL, Thio CL, Martin MP, et al. Genetic variation in IL28B and spontaneous clearance of hepatitis C virus. Nature 2009; 461: 798–801.

50. Young AM, Crosby RA, Oser CB, et al. Hepatitis C viremia and genotype distribution among a sample of nonmedical prescription drug users exposed to HCV in rural Appalachia. J Med Virol 2012; 84: 1376–1387.

51. Smith DJ, Combellick J, Jordan AE, et al. Hepatitis C virus (HCV) disease progression in people who inject drugs (PWID): A systematic review and meta-analysis. Int J Drug Policy 2015; 26: 911–921.

52. Erman A, Krahn MD, Hansen T, et al. Estimation of fibrosis progression rates for chronic hepatitis C: a systematic review and meta-analysis update. BMJ Open 2019; 9: e027491.

53. Aponte-Meléndez Y, Mateu-Gelabert P, Eckhardt B, et al. Hepatitis C virus care cascade among people who inject drugs in Puerto Rico: Minimal HCV treatment and substantial barriers to HCV care. Drug Alcohol Depend Rep 2023; 8: 100178.

54. Bull-Otterson L, Huang Y-LA, Zhu W, et al. Human Immunodeficiency Virus and Hepatitis C Virus Infection Testing Among Commercially Insured Persons Who Inject Drugs, United States, 2010-2017. J Infect Dis 2020; 222: 940–947.

55. Thompson WW, Symum H, Sandul A, et al. Vital Signs: Hepatitis C Treatment Among Insured Adults - United States, 2019-2020. MMWR Morb Mortal Wkly Rep 2022; 71: 1011–1017.

56. Kapadia SN, Zhang H, Gonzalez CJ, et al. Hepatitis C Treatment Initiation Among US Medicaid Enrollees. JAMA Netw Open 2023; 6: e2327326.

57. Berg T, Naumann U, Stoehr A, et al. Real-world effectiveness and safety of glecaprevir/pibrentasvir for the treatment of chronic hepatitis C infection: data from the German Hepatitis C-Registry. Aliment Pharmacol Ther 2019; 49: 1052–1059.

58. Cornberg M, Stoehr A, Naumann U, et al. Real-World Safety, Effectiveness, and Patient-Reported Outcomes in Patients with Chronic Hepatitis C Virus Infection Treated with Glecaprevir/Pibrentasvir: Updated Data from the German Hepatitis C-Registry (DHC-R). Viruses 2022; 14: 1541.

59. Sowah LA, Smeaton L, Brates I, et al. Perspectives on Adherence From the ACTG 5360 MINMON Trial: A Minimum Monitoring Approach With 12 Weeks of Sofosbuvir/Velpatasvir in Chronic Hepatitis C Treatment. Clin Infect Dis Off Publ Infect Dis Soc Am 2023; 76: 1959– 1968.

60. Cunningham EB, Amin J, Feld JJ, et al. Adherence to sofosbuvir and velpatasvir among people with chronic HCV infection and recent injection drug use: The SIMPLIFY study. Int J Drug Policy 2018; 62: 14–23.

61. Turner BJ, Hecht FM, Ismail RB. CD4+ T-lymphocyte measures in the treatment of individuals infected with human immunodeficiency virus type 1. A review for clinical practitioners. Arch Intern Med 1994; 154: 1561–1573.

62. Gopalappa C, Farnham PG, Chen Y-H, et al. Progression and Transmission of HIV/AIDS (PATH 2.0). Med Decis Mak Int J Soc Med Decis Mak 2017; 37: 224–233.

63. Singh S, France AM, Chen Y-H, et al. Progression and Transmission of HIV (PATH 4.0) – A new Agent-based Evolving Network Simulation for Modeling HIV Transmission Clusters. Math Biosci Eng MBE 2021; 18: 2150–2181.

64. Rodríguez B, Sethi AK, Cheruvu VK, et al. Predictive Value of Plasma HIV RNA Level on Rate of CD4 T-Cell Decline in Untreated HIV Infection. JAMA 2006; 296: 1498–1506.

65. Gras L, Kesselring AM, Griffin JT, et al. CD4 cell counts of 800 cells/mm3 or greater after 7 years of highly active antiretroviral therapy are feasible in most patients starting with 350 cells/mm3 or greater. J Acquir Immune Defic Syndr 1999 2007; 45: 183–192.

66. Monitoring Selected National HIV Prevention and Care Objectives by Using HIV Surveillance Data—United States and 6 Dependent Areas, 2019. 26.

67. Wang L, Krebs E, Min JE, et al. Combined estimation of disease progression and retention on antiretroviral therapy among treated individuals with HIV in the USA: a modelling study. Lancet HIV 2019; 6: e531–e539.

68. Hanmer J, Lawrence WF, Anderson JP, et al. Report of nationally representative values for the noninstitutionalized US adult population for 7 health-related quality-of-life scores. Med Decis Mak Int J Soc Med Decis Mak 2006; 26: 391–400.

69. Pyne JM, French M, McCollister K, et al. Preference-weighted health-related quality of life measures and substance use disorder severity. Addict Abingdon Engl 2008; 103: 1320– 1329; discussion 1330-1332.

70. Saeed YA, Phoon A, Bielecki JM, et al. A Systematic Review and Meta-Analysis of Health Utilities in Patients With Chronic Hepatitis C. Value Health J Int Soc Pharmacoeconomics Outcomes Res 2020; 23: 127–137.

71. Tran BX, Nguyen LH, Ohinmaa A, et al. Longitudinal and cross sectional assessments of health utility in adults with HIV/AIDS: a systematic review and meta-analysis. BMC Health Serv Res 2015; 15: 7.

72. Aden B, Dunning A, Nosyk B, et al. Impact of Illicit Drug Use on Health-Related Quality of Life in Opioid Dependent Patients Undergoing HIV Treatment. J Acquir Immune Defic Syndr 1999 2015; 70: 304–310.

73. Nosyk B, Min JE, Krebs E, et al. The Cost-Effectiveness of Human Immunodeficiency Virus Testing and Treatment Engagement Initiatives in British Columbia, Canada: 2011–2013. Clin Infect Dis 2018; 66: 765–777.

74. Statnet Development Team (Pavel N. Krivitsky, Mark S. Handcock, David R. Hunter, Carter T. Butts, Chad Klumb, Steven M. Goodreau, and Martina Morris). Statnet: Tools for the Statistical Modeling of Network Data, http://statnet.org (2003).

75. Cohen MS, Chen YQ, McCauley M, et al. Prevention of HIV-1 Infection with Early Antiretroviral Therapy. N Engl J Med 2011; 365: 493–505.

76. Aspinall EJ, Nambiar D, Goldberg DJ, et al. Are needle and syringe programmes associated with a reduction in HIV transmission among people who inject drugs: a systematic review and meta-analysis. Int J Epidemiol 2014; 43: 235–248.

77. Centers for Disease Control and Prevention. Effective HIV Prevention Strategies, https://www.cdc.gov/hiv/risk/estimates/preventionstrategies.html (2022, accessed 19 August 2024).

78. Stout NK, Goldie SJ. Keeping the Noise Down: Common Random Numbers for Disease Simulation Modeling. Health Care Manag Sci 2008; 11: 399–406.

79. Hajarizadeh B, Cunningham EB, Valerio H, et al. Hepatitis C reinfection after successful antiviral treatment among people who inject drugs: A meta-analysis. J Hepatol 2020; 72: 643–657.

80. Esmaeili A, Mirzazadeh A, Carter GM, et al. Higher incidence of HCV in females compared to males who inject drugs: A systematic review and meta-analysis. J Viral Hepat 2017; 24: 117–127.

81. Valencia J, Alvaro-Meca A, Troya J, et al. High rates of early HCV reinfection after DAA treatment in people with recent drug use attended at mobile harm reduction units. Int J Drug Policy 2019; 72: 181–188.

82. Caven M, Malaguti A, Robinson E, et al. Impact of Hepatitis C treatment on behavioural change in relation to drug use in people who inject drugs: A systematic review. Int J Drug Policy 2019; 72: 169–176.

83. Coyle C, Moorman AC, Bartholomew T, et al. The HCV care continuum: linkage to HCV care and treatment among patients at an urban health network, Philadelphia, PA. Hepatol Baltim Md 2019; 70: 476–486.

84. Kochanek KD. Mortality in the United States, 2022.

85. How Are Opioid Settlement Funds Being Spent So Far? | Health Affairs, https://www.healthaffairs.org/content/forefront/opioid-settlement-funds-being-spent-so-far (accessed 11 June 2024).

86. Issue Brief: The Role of Housing in Ending the HIV Epidemic | Policy and Law | HIV/AIDS | CDC, https://www.cdc.gov/hiv/policies/data/role-of-housing-in-ending-the-hiv-epidemic.html (2024, accessed 11 June 2024).

87. Barocas JA, Nall SK, Axelrath S, et al. Population-Level Health Effects of Involuntary Displacement of People Experiencing Unsheltered Homelessness Who Inject Drugs in US Cities. JAMA 2023; 329: 1478–1486.

88. Castrucci BC, Auerbach J. Meeting Individual Social Needs Falls Short Of Addressing Social Determinants Of Health, https://www.healthaffairs.org/do/10.1377/forefront.20190115.234942/full/ (accessed 26 September 2024).

89. Tempalski B, Pouget ER, Cleland CM, et al. Trends in the Population Prevalence of People Who Inject Drugs in US Metropolitan Areas 1992–2007. PLOS ONE 2013; 8: e64789.

90. Spiller MW, Gile KJ, Handcock MS, et al. Evaluating Variance Estimators for Respondent-Driven Sampling. J Surv Stat Methodol 2018; 6: 23–45.

91. Zhu L, Menzies NA, Wang J, et al. Estimation and correction of bias in network simulations based on respondent-driven sampling data. Sci Rep 2020; 10: 6348.

92. Tanz LJ. Routes of Drug Use Among Drug Overdose Deaths — United States, 2020–2022. MMWR Morb Mortal Wkly Rep; 73. Epub ahead of print 2024. DOI: 10.15585/mmwr.mm7306a2.

93. Ciccarone D, Holm N, Ondocsin J, et al. Innovation and adaptation: The rise of a fentanyl smoking culture in San Francisco. PLOS ONE 2024; 19: e0303403.

94. Petitti DB, Lin JS, Owens DK, et al. Collaborative Modeling: Experience of the U.S. Preventive Services Task Force. Am J Prev Med 2018; 54: S53–S62.

