## Supplement for "Strategies to Achieve HIV and HCV Infection Incidence Targets Among People Who Inject Drugs: A Stochastic Network-Based Multi-Disease Transmission Modeling Study"

### Parameter Estimation

#### Injection Drug Use Dynamics

##### *Injecting Initiation Rate*

Individuals enter our simulation when they initiate injection drug use (IDU). We assumed all individuals initiating IDU were aged 18–24 years, HIV negative, and HCV negative.[1–3] We calibrated the number of individuals initiating IDU per month to yield a 25% increase in the simulated people who inject drugs (PWID) population size over a period of 10 years in the baseline scenario (Figure S1). The target of 25% reflected a slowing of the rate of increase in prevalence of IDU compared to that observed in the 2010s.[4–6]

*Figure S1. Size of the Modeled Population.*

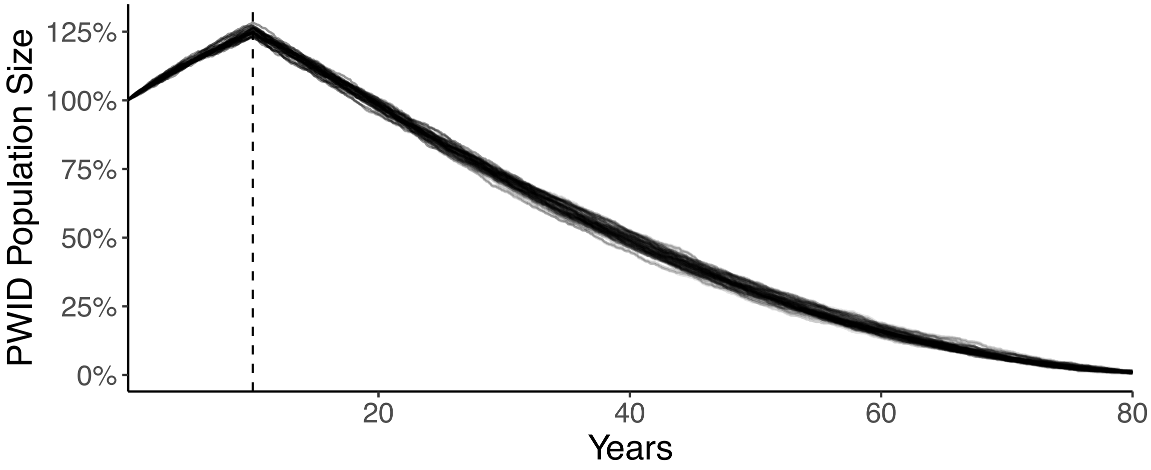

Note: Interventions stop and population is closed after 10 years (dashed line).

##### *Injecting Cessation Rate, Relapse Rate, and Permanent Cessation*

We estimated monthly drug injection cessation rates, relapse rates, and the probability of permanent drug injection cessation from longitudinal data collected from the ALIVE Study cohort.[7] Trajectories from the ALIVE Study are largely consistent with those observed among a cohort of young PWID in San Francisco (UFO Study) and PWID in Vancouver

(Vancouver Injection Drug Users Study).[8,9] Individuals were considered to be "temporary" former PWID if they did not inject drugs during the monthly timestep but would eventually resume IDU in a future timestep.  Individuals were considered to be "permanent" former PWID if they did not inject drugs during the monthly timestep and would never resume using injection drugs. Figure S2 depicts the prevalence and population size of modeled current IDU, temporary drug injection cessation, and permanent drug injection cessation states over time. Figure S3 shows examples of individual-level transitions between states.

*Figure S2. Baseline scenario prevalence (top) and population size (bottom) of current IDU, temporary drug injection cessation, and permanent drug injection cessation.*

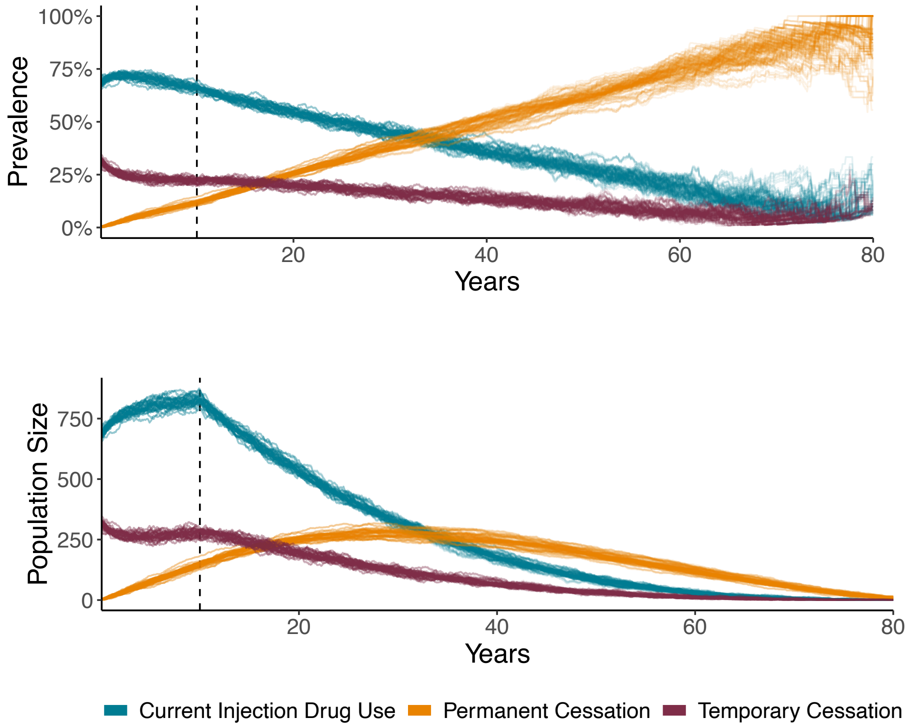

Note: Interventions stop and population is closed after 10 years (dashed line).

*Figure S3. Sample of baseline scenario individual-level trajectories of injection drug use. Trajectories end at death.*

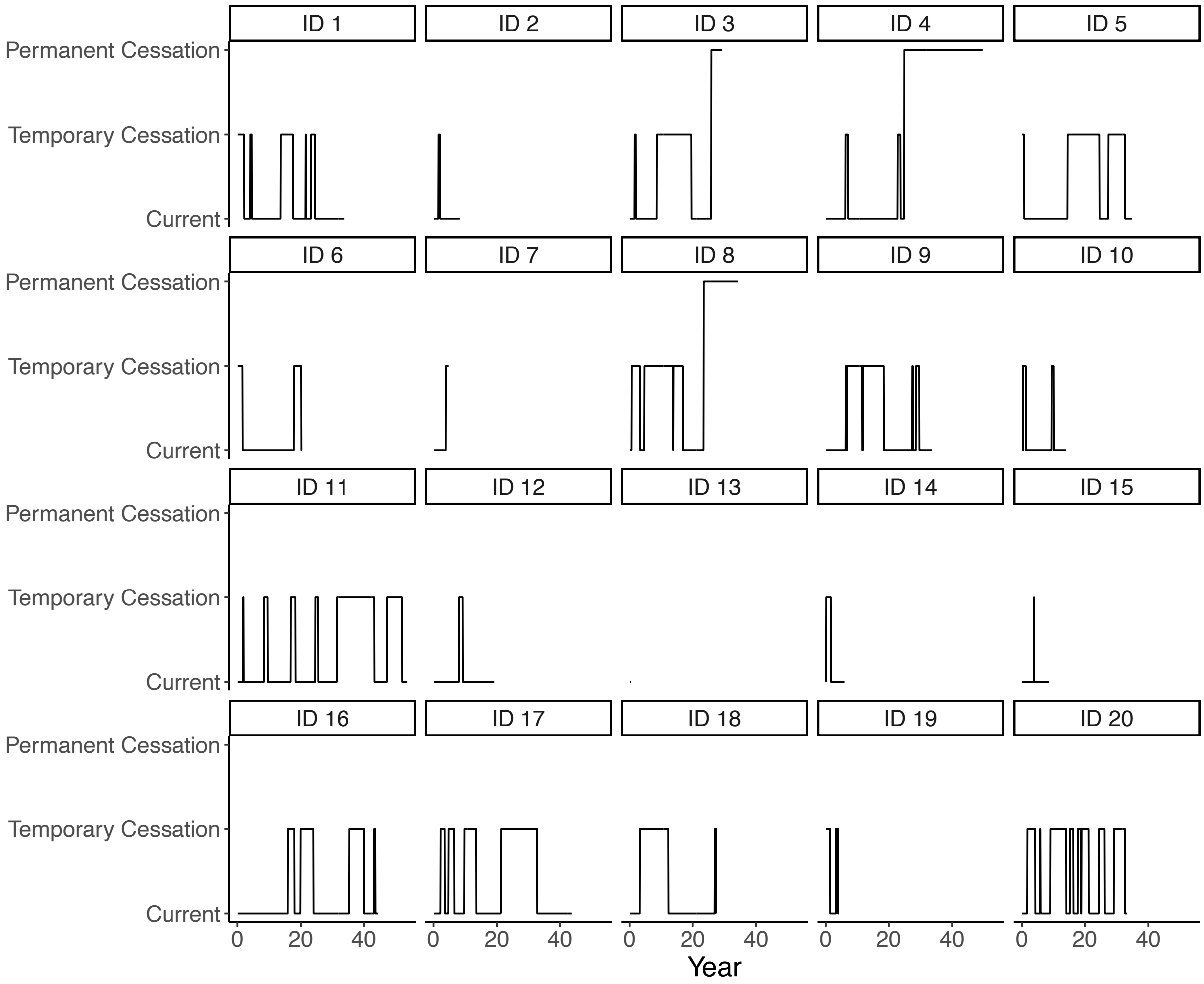

#### *Consistent Population Size for Per-Person Calculations*

We defined a consistent population size (n = 1240) to compute per-person life-years and quality-adjusted life-years. This consistent population size was based on the size of the simulated population at simulation start (n = 1000), plus the number of PWID initiating and entering the simulated population during the ten-year intervention time-horizon (n = 480), proportionally weighted by the duration of potential exposure to the intervention.

#### Mortality

##### *Excess Mortality Due to Injection Drug Use*

We estimated monthly rates of mortality due to current or former IDU based on published data from a cohort of treated, opioid-dependent individuals.[10] We extracted the standardized mortality ratios (SMRs) for former drug use (1.8) and current drug use (6.1). Standardized mortality ratios for this cohort decreased with increasing age. Although age-specific data were not reported separately by current and former drug use status, inclusion criteria and length of follow-up for the cohort suggested that different proportions of current and former by age did not explain the observed age pattern. Further, a global systematic review and a separate analysis of data from the ALIVE cohort both reported decreasing SMRs with increasing age.[11,12] Following this evidence, applying a fixed SMR to age-specific background mortality would likely overestimate the rate of death due to drug use among older PWID. Therefore, we split age-specific SMRs from Evans et al. (2015) into age-specific current and former SMRs using the ratio of current and former to overall SMRs. We then interpolated and extrapolated these SMRs across the full range of ages included in the model (18 to 100) by fitting a linear model to predict log-transformed SMRs, offset by 1. This specification generated smooth, monotonically decreasing, age-specific SMRs, that asymptote at 1 (Figure S4). We applied these age-specific SMRs to age-specific mortality rates from the 2019 United States life table[13] to generate age-specific excess mortality rates due to current and former drug use (Figure S5).

*Figure S4. Modeled age-specific standardized mortality ratios for people who currently or formerly (temporary or permanent) injected drugs.*

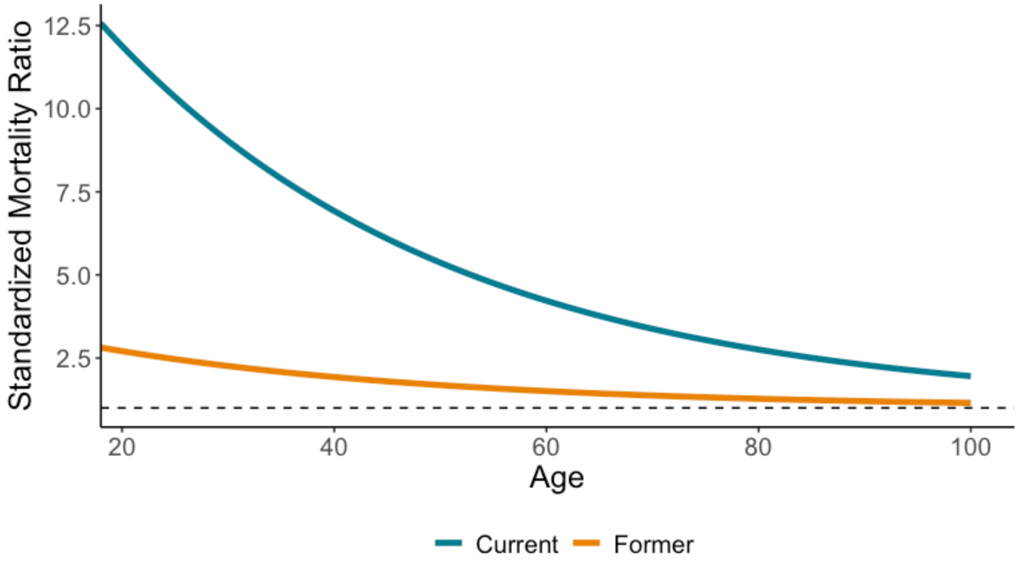

*Figure S5. Modeled age-specific monthly excess mortality rates for people who currently and formerly (temporary or permanent) injected drugs. Age is capped at 80 years for visualization purposes.*

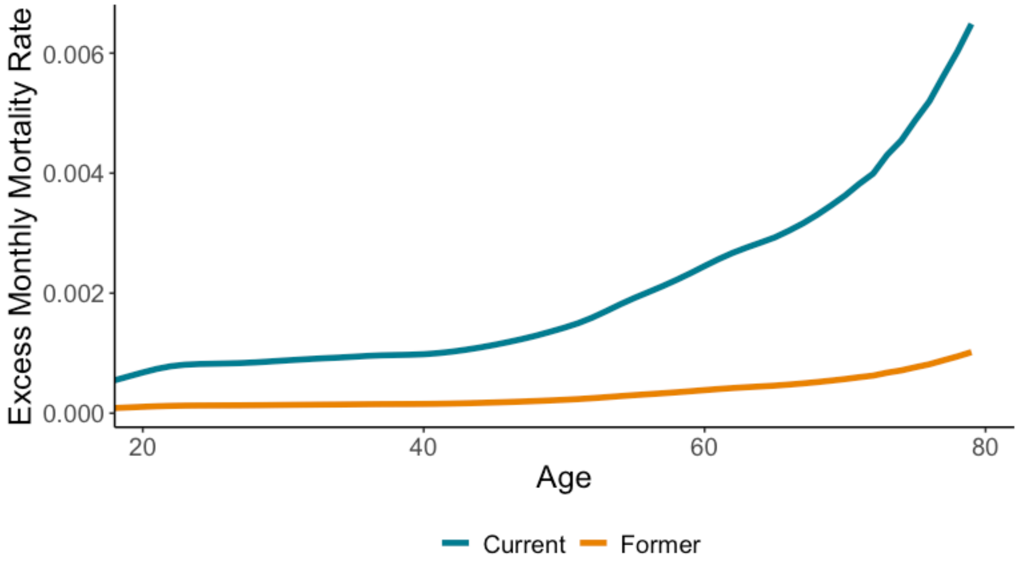

##### *Excess Mortality Due to HCV Infection*

We included excess mortality due to HCV infection among individuals with compensated (METAVIR F4) or decompensated cirrhosis. We computed liver-related mortality rates from a cohort of patients with cirrhosis.[14] We included a mortality risk reduction for individuals in our model who had cirrhosis and were treated with direct-acting antivirals to reach SVR. We used a hazard ratio of 0.29 for the reduction in liver-related mortality rates among those with cirrhosis achieving SVR compared to those not achieving SVR.[15]

##### *Excess Mortality Due to HIV Infection*

We included excess mortality due to HIV as a continuous function of CD4 count (Figure S6). We leveraged the relationship between CD4 count and mortality estimated in an evidence synthesis of seroconverter cohorts.[16] The HIV-related mortality rate increased sharply at CD4 counts below 200 cells per microliter. Our approach to modeling HIV-related mortality entirely encoded the benefits of antiretroviral treatment (ART) through changes in CD4 count.

*Figure S6. Estimated annual HIV-related mortality rates by CD4 count. CD4 count is capped at 10 and 800 cells per microliter for visualization purposes.*

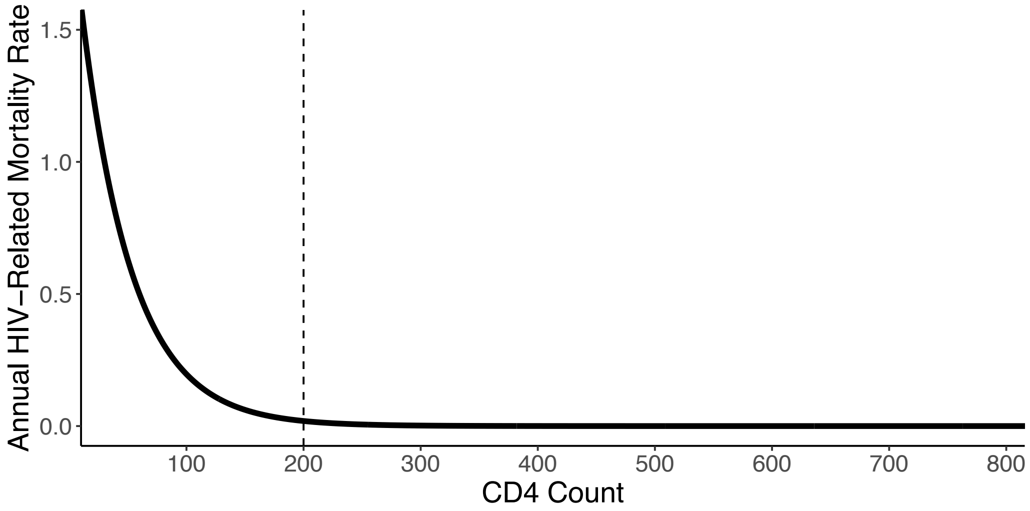

#### HIV and HCV Infection Treatment and Progression

Persons with HCV infection faced monthly fibrosis stage-specific probabilities of progression (Figure S7).[17,18] Progression was halted, but not reversed, when individuals reached SVR. In our analysis, individuals with HIV/HCV co-infection progressed faster than individuals with HCV mono-infection.[19] Individuals with HIV/HCV co-infection on ART had a progression rate ratio of 1.7 compared to individuals with HCV mono-infection, while individuals with HIV/HCV co-infection that were not on ART had a progression rate ratio of 2.5.[20]

We implemented a simplified HCV care cascade. In our baseline analysis, persons with HCV infection had a probability of 0.008 for being tested for HCV each month, 23% of individuals tested were successfully linked to care and began treatment, 96% completed treatment, and 94% reached SVR after completing treatment.[1,21–29] Observed care cascades are highly variable across settings. To isolate the effects of treatment, we did not include additional risk reduction for reinfection through behavior change following treatment. In a sensitivity analysis, we evaluated the impact of a relative risk of 0.34 for reinfection after treatment and a lower treatment completion percentage (75%).[30–34]

*Figure S7. Cumulative duration of HCV infection prior to fibrosis progression.*

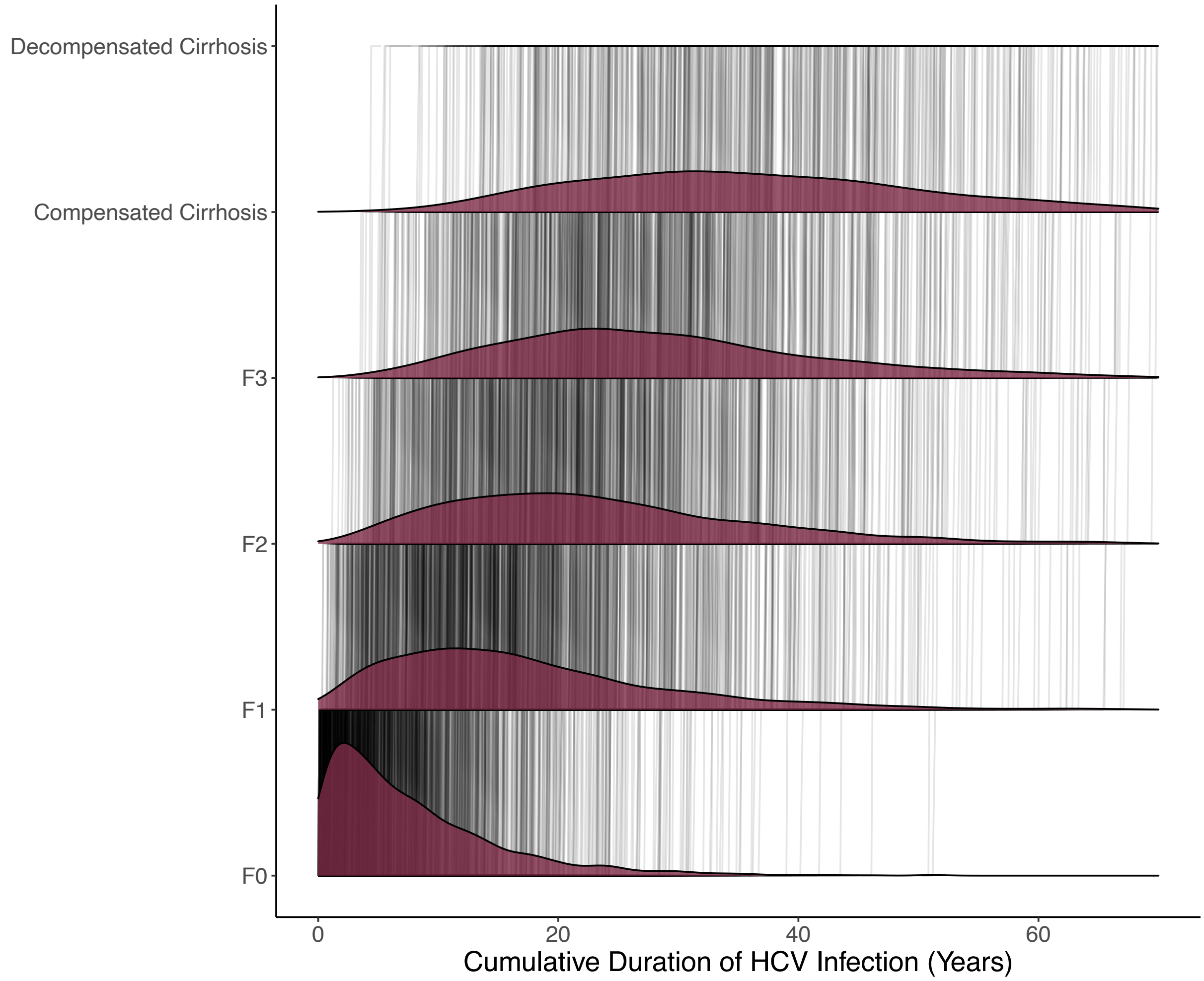

Individuals with HIV that were not on treatment faced a constant monthly decline in their CD4 count (-4.7 per month). When individuals start ART (for the first time or after a treatment interruption), they experienced the greatest increase in CD4 count (+22.7 per month) for the first six months, followed by a slower increase (+13.3 per month) during months seven to 36. We capped CD4 count levels at a threshold that depended on the individuals' CD4 nadir (Table S1).

*Table S1. CD4 count ceilings as a function of CD4 count nadirs.*

| Nadir CD4 | CD4 Ceiling |
|---|---|
| Less than 50 | 410 |
| 50-200 | 548 |
| 201-350 | 660 |
| 351-500 | 780 |
| Greater than 500 | 870 |

Additionally, when individuals experienced a treatment interruption, we assumed their CD4 count immediately declined to their nadir. A study of HIV-positive PWID in Vancouver found a high degree of "churn," with HIV-positive PWID frequently cycling on and off ART.[35] Figure S8 depicts a sample of modeled CD4 trajectories among PWID with HIV. In our model, ever-treated PWID whose cause of death was HIV lived for an average of ten years longer than never-treated PWID. Ever-treated PWID who were on ART for a majority of their duration of HIV infection lived for more than 20 years longer than never-treated PWID.

*Figure S8. Sample of baseline scenario individual-level CD4 trajectories. Trajectories end at death (regardless of cause of death).*

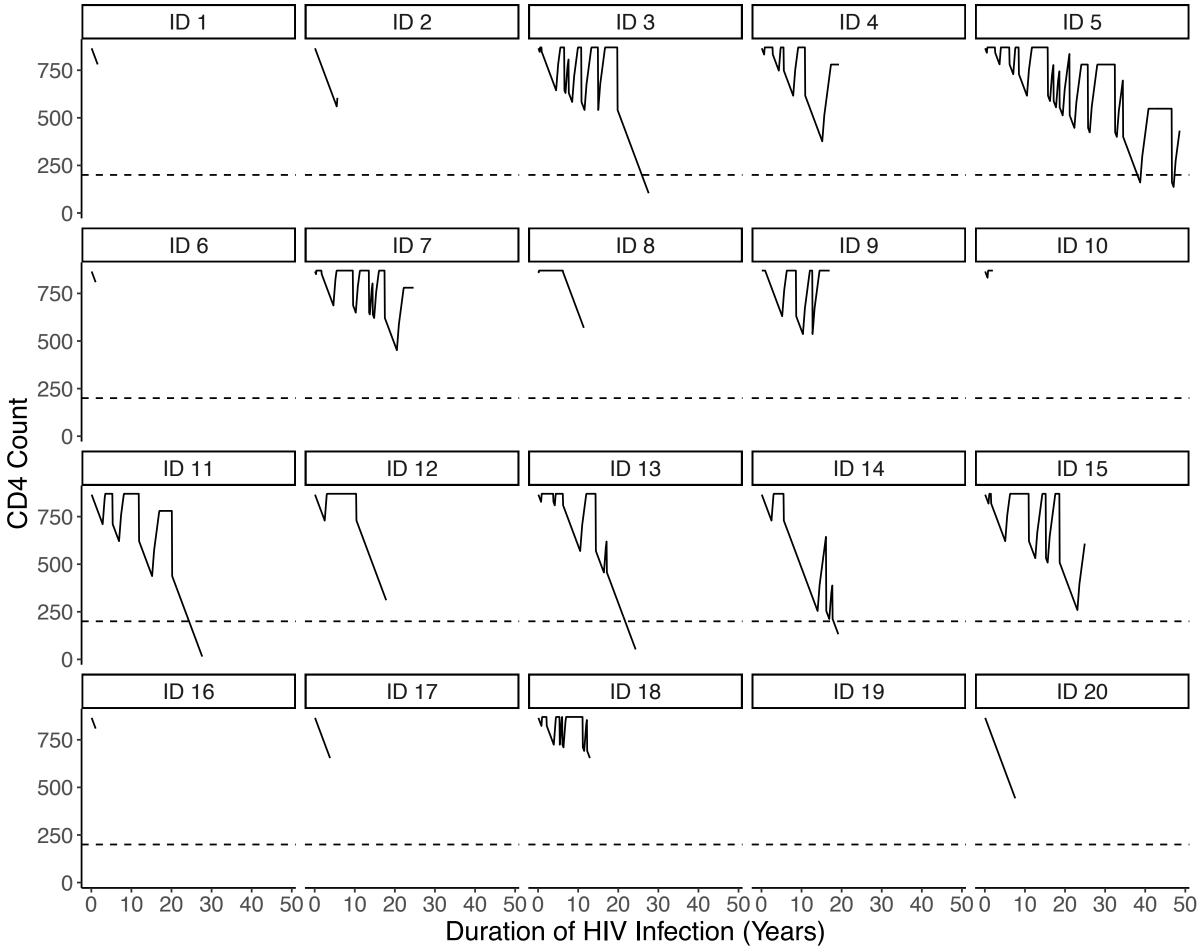

#### Health Utilities

We assume health utilities are multiplicative and included utilities for age group, current or former PWID status, HCV, and HIV. We adjust published utilities related to HIV and HCV to avoid double counting of utility decrements.[36,37] Specifically, we believed that correlations between age, PWID status (current/former), HIV prevalence, and HCV prevalence prevented us from interpreting published utilities as independent. For HCV, we assumed that individuals who reached SVR and were in METAVIR stages F0-F3 did not experience a health utility decrement due to HCV (e.g., their HCV-related utility was 1). Using F0-F3 with SVR as the reference category, we adjusted published utilities from Saeed et al. (2020) (Table S2).

*Table S2. Approach to adjusting published HCV-related health utilities.*

| Category | Published[36] | Adjusted | Method |
|---|---|---|---|
| F0-F3 with SVR | 0.786 | 1 | Multiply by 1/0.786 |
| F0-F3 without SVR | 0.751 | 0.955 | Multiply by 1/0.786 |
| F4 with SVR | 0.671 | 0.958 | Scale adjusted disutility (1-(0.671/0.786)) by mortality HR (0.29) |
| F4 without SVR | 0.671 | 0.854 | Multiply by 1/0.786 |
| Decompensated with SVR | 0.602 | 0.932 | Scale adjusted disutility (1-(0.602/0.786)) by mortality HR (0.29) |
| Decompensated without SVR | 0.602 | 0.766 | Multiply by 1/0.786 |

For HIV, we assumed that individuals with CD4 counts of at least 500 cells per microliter did not experience a health utility decrement due to HIV. We also estimated that individuals with symptomatic HIV or AIDS, who were on ART, had utilities that were 13% higher than individuals who are not on ART.[38–40] Using CD4 ≥ 500 as the reference category, we adjusted published utilities from Tran et al. (2015) (Table S3).

*Table S3. Approach to adjusting published HIV-related health utilities.*

| Category | Published[37] | Adjusted | Method |
|---|---|---|---|
| CD4 ≥ 500 | 0.861 | 1 | Multiply by 1/0.861 |
| 500 > CD4 ≥ 200, ART | 0.844 | 0.980 | Multiply by 1/0.861 |
| 500 > CD4 ≥ 200, No ART | - | 0.867 | Multiply adjusted with ART by 1/1.13 |
| 200 ≥ CD4, ART | 0.688 | 0.799 | Multiply by 1/0.861 |
| 200 ≥ CD4, No ART | - | 0.707 | Multiply adjusted with ART by 1/1.13 |

Using CD4 count-specific utilities from Tran et al. (2015), that were for individuals not on ART as the reference, resulted in differences of less than 0.02 across all categories.

#### Network Simulations

We fit separable temporal exponential-family random graph models (STERGMs) to simulate networks of injection equipment sharing partnerships and sexual partnerships. STERGMs are dynamic extensions of exponential-family random graph models (ERGMs), which describe the probability of a network as a function of covariates. Covariates can include node-level, dyad-level, and network-level attributes. A general ERGM is written as:

$$P(Y = y) = \frac{\exp(\theta^T g(y))}{k(\theta)}$$

where, $y$ is one realization of a random variable for the state of the network ($Y$), $g(y)$ is a vector of covariates, $\theta$ is a vector of coefficients for the covariates, and $k(\theta)$ is a normalizing constant defined as the sum of the numerator over all possible networks $y$. The models were fit using a well-supported suite of packages developed in R by the Statnet project.[41] Coefficients ($\theta$) were estimated through a Markov Chain Monte Carlo process, comparing simulated network structure statistics to observed network structure statistics. Once the model coefficients were estimated, they were used to simulate new networks.

We analyzed the 2018 NHBS PWID data to estimate the observed network structure statistics that were used as targets to fit STERGMs (Table S4).[2] For each of the 23 NHBS project areas, we computed network structure target statistics as the mean across respondents from the project area. We then fit our network models to the median value over the 23 project areas. This approach allowed us to maintain confidentiality of NHBS data. For some indicators, we derived measures from the "last partner" series of questions. The responses to these questions were based on characteristics of either the last equipment sharing or last sexual partners. As a result, we assumed that individuals' last partners were representative of all partners.

Based on our analysis of 2018 NHBS PWID data, 21% of respondents' last equipment sharing partners were also sexual partners.[2] We used this to inform overlap between the sexual and equipment sharing networks at the partnership level. To ensure that our simulated networks maintained the observed percentage of dual partnerships, we included the equipment sharing network as a dyad-level covariate for sexual partnership formation, and we included the sexual network as a dyad-level covariate for equipment sharing partnership formation. Additionally, 54% of respondents' last sexual partners were "probably" or "definitely" PWID.[2] We used this to inform overlap between the sexual and equipment sharing networks at the network level. Specifically, we used this percentage to reduce the number of modeled sexual ties within our PWID network, and then estimated out-of-network partnerships with non-PWID sexual partners.

*Table S4. Target statistics, reflecting the median value across 23 project areas, derived from analysis of 2018 NHBS data among people who inject drugs.*[2]

| **Target Statistic** | **Value** | **Question Series** |
|---|---|---|
| Mean number of equipment sharing partnerships (last 12 months) | 4.5 | General |
| Mean number of sexual partnerships (last 12 months) | 6.5 | General |
| Percentage of PWID with no equipment sharing partners (past 12 months) | 35% | General |
| Percentage of PWID with no sexual partners (last 12 months) | 15% | General |
| Equipment sharing partnerships that were also sexual partnerships | 21% | Last Partner |
| Sexual partners that were "probably" or "definitely" PWID | 54% | Last Partner |
| Mean sexual partnership duration (years) | 4 | Last Partner |

Mean equipment sharing partnership duration was the only network target statistic that did not come from the 2018 NHBS PWID data, due to unavailability of this measure. We assumed a mean duration of three years, consistent with other PWID network models.[42]

We confirmed that our simulated networks remained close to target statistics as they dynamically evolved (Figure S9).

*Figure S9. Mean degree of equipment sharing and sexual partnerships during the 10-year intervention period in the baseline scenario.*

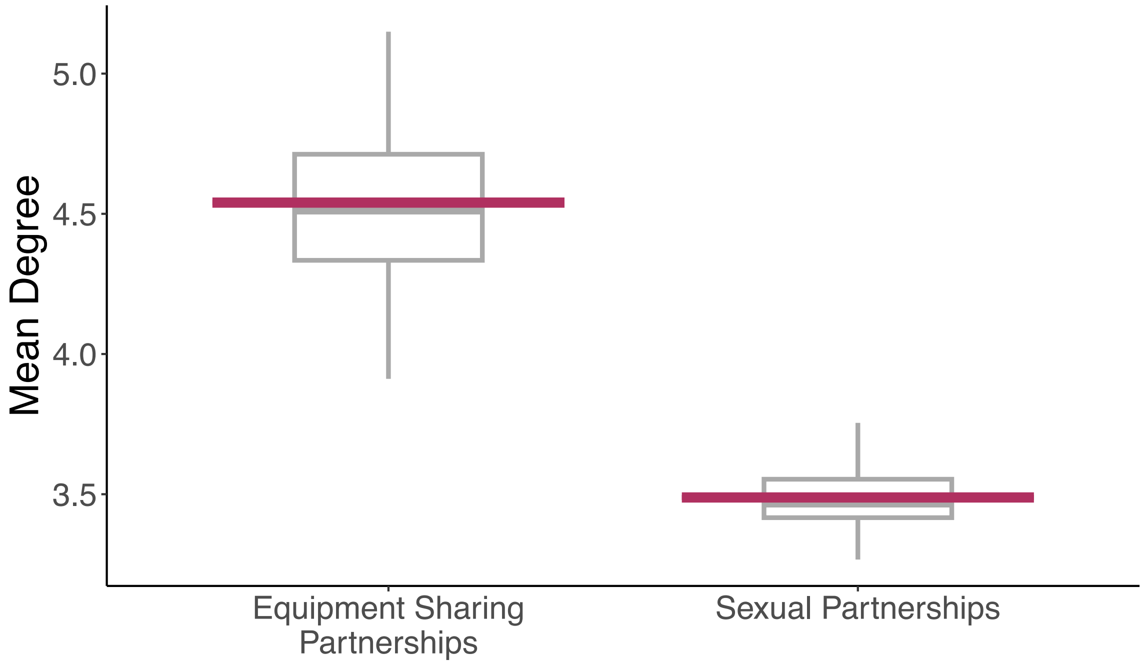

#### Calibration

We calibrated monthly probabilities of HCV and HIV transmission. We used HIV (8.6%) and HCV (44.1%) infection prevalence as calibration targets.[43] We assumed steady prevalence in our baseline scenario. To reduce the parameter sample space and inform our prior distributions, we imposed structure on the relationship between HCV and HIV transmission through equipment sharing, and HIV transmission through equipment sharing versus sexual activity. Specifically, we assumed that the probability of HIV transmission via equipment sharing was lower than the probability of HCV transmission via equipment sharing, consistent with literature demonstrating higher per-contact infectivity of HCV and greater environmental survival of HCV.[44–46] Additionally, we assumed that the probability of HIV transmission via sexual activity was lower than the probability of HIV transmission via equipment sharing, based on the higher prevalence of condomless vaginal sex compared to condomless anal sex among 2018 NHBS PWID respondents.[2] We used a grid search to calibrate transmission probabilities (Table S5). For each parameter set, we computed the mean absolute percentage error (MAPE) between the model estimates and calibration targets. We defined the best-fitting parameter set as the set that minimized the MAPE. We found that this parameter set also minimized sum of squared errors. Due to the computational intensity of our agent-based network model, we used the single best-fitting parameter set for our main analyses and explored the impacts of alternative parameters through one-way sensitivity analyses (Figure S10 and S11). As a result, uncertainty in our main analysis is entirely stochastic uncertainty, and does not include parameter uncertainty.

*Table S5. Prior distributions of parameters used to define transmission probabilities.*

| **Calibrated Parameter** | **Prior** |
|---|---|
| Monthly probability of HCV transmission | Unif(0.004, 0.01) |
| Relative infectiousness of HCV transmission vs. HIV transmission through equipment sharing | Unif(1.0, 2.0) |
| Relative infectiousness of HIV transmission through equipment sharing vs. sexual activity | Unif(2, 5) |

*Figure S10. Estimates of HCV infection prevalence from the calibrated parameter prior distribution and selected parameter set, across five stochastic runs.*

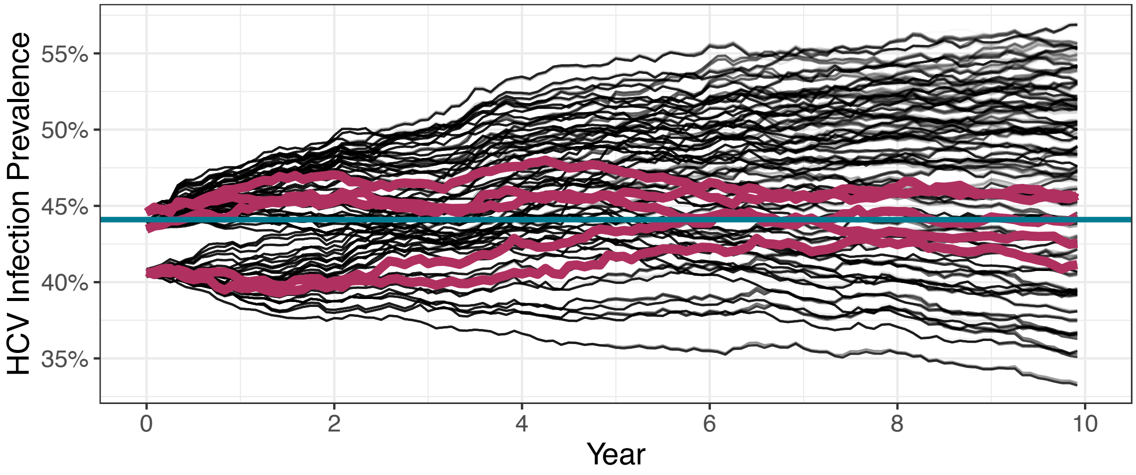

Note: Horizontal blue line is the calibration target. Maroon lines are from the selected parameter set.

*Figure S11. Estimates of HIV infection prevalence from the calibrated parameter prior distribution and selected parameter set, across five stochastic runs.*

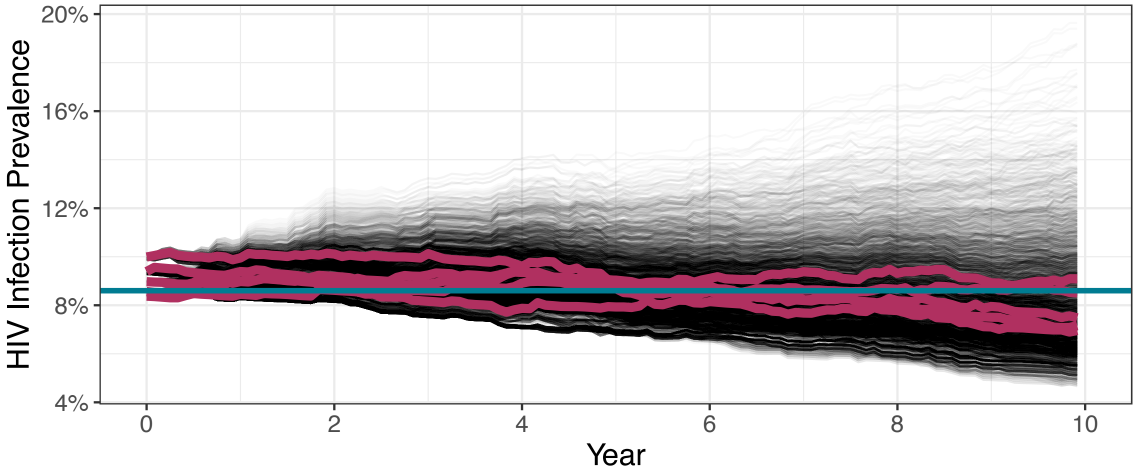

Note: Horizontal blue line is the calibration target. Maroon lines are from the selected parameter set.

#### Common Random Numbers for Stochastic Noise Reduction

We developed and implemented methods to leverage common random numbers to reduce stochastic noise in our agent-based transmission dynamic model. There were 28 stochastic events in our model. Examples of stochastic events included infection transmission, testing, and treatment, whether an individual was using harm reduction services at a given timepoint, and mortality. Traditionally, stochastic models are run many times to generate precise estimates of summary measures. Agent-based network models are more computationally intensive than compartmental or microsimulation models that do not include agent interactions. As a result, the standard approach to reduce stochastic noise through repeated simulation can result in substantial computational burden.

Common random numbers enable use of the same random draw for a stochastic event in different scenarios.[47] This allows us to estimate precise intervention effects with fewer simulation runs. Use of common random numbers for all stochastic events eliminates stochastic noise when computing intervention effects, but such exhaustive implementation can also pose a computational challenge. We implemented common random numbers for all stochastic events except partnership formation and termination for the sexual and equipment sharing networks, and we implemented common random numbers for transmission events at the individual-level as opposed to at the dyad-level. As a result, we substantially reduced, but did not entirely eliminate, the effects of stochastic noise. Nonetheless, these methods enabled us to explore far more intervention scenarios than would otherwise be computationally feasible.

When implementing common random numbers, we assumed that conditional stochastic events (e.g., treatment is conditional on testing positive and testing positive is conditional on being infected) were indexed on the event number, not on the timestep during which the event occurred. In doing so, interventions with a beneficial effect always resulted in benefits with respect to direct effects (although increases or decreases in outcomes are possible as a result of indirect effects).

Finally, although we aimed to reduce stochastic noise when comparing outcomes between intervention scenarios, we desired to capture the range of effects due to stochastic uncertainty within a scenario. In other words, we simulated many stochastic “states of the world,” but held these “states of the world” as constant as possible when conducting paired comparisons between baseline and intervention scenarios. For this analysis, we found that sampling 150 “states of the world” produced sufficiently precise estimates of intervention effects on incidence of HIV and HCV infections (Figures S12 and S13).

*Figure S12. Convergence of estimates of the percentage change in HIV incidence between baseline and intervention scenarios.*

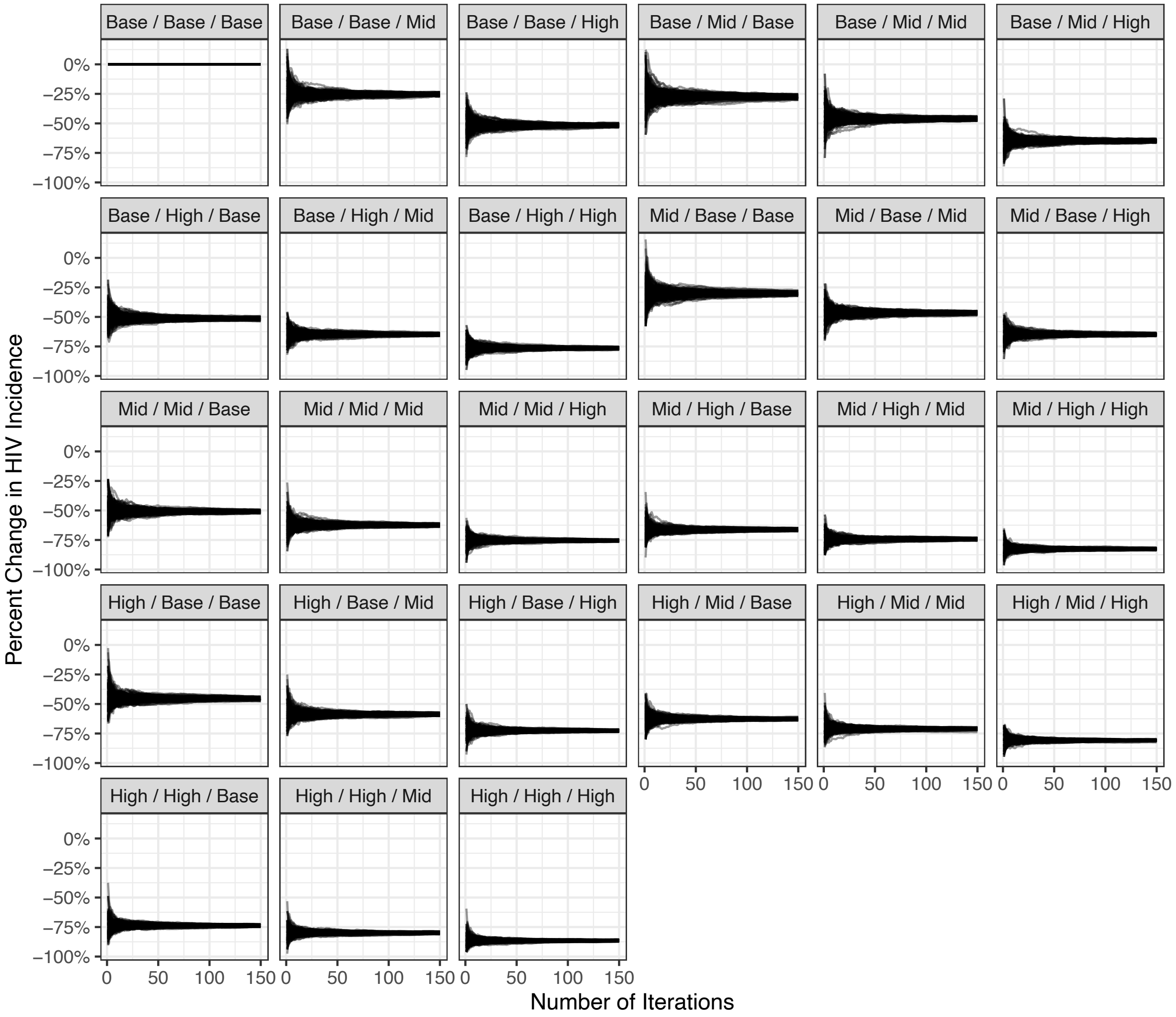

Footnote: Panel labels report cessation level first, prevention intervention level second, and test and treat level third. Baseline cessation rate is 16 per 100 person-years, moderate cessation rate is 31 per 100 person-years, and high cessation rate is 46 per 100 person-years. Baseline prevention intervention coverage for injection equipment sharing is 53%, moderate coverage is 68%, and high coverage is 83%. Baseline prevention intervention coverage for sex is 20%, moderate coverage is 35%, and high coverage is 50%. The percentage of people with HCV infection reaching SVR per year is 3% at baseline test and treat, 16% at moderate test and treat, and 28% at high test and treat. The percentage of people with HIV that are virally suppressed is 44% at baseline test and treat, 58% at moderate test and treat, and 71% at high test and treat.

*Figure S13. Convergence of estimates of the percentage change in HCV incidence between baseline and intervention scenarios.*

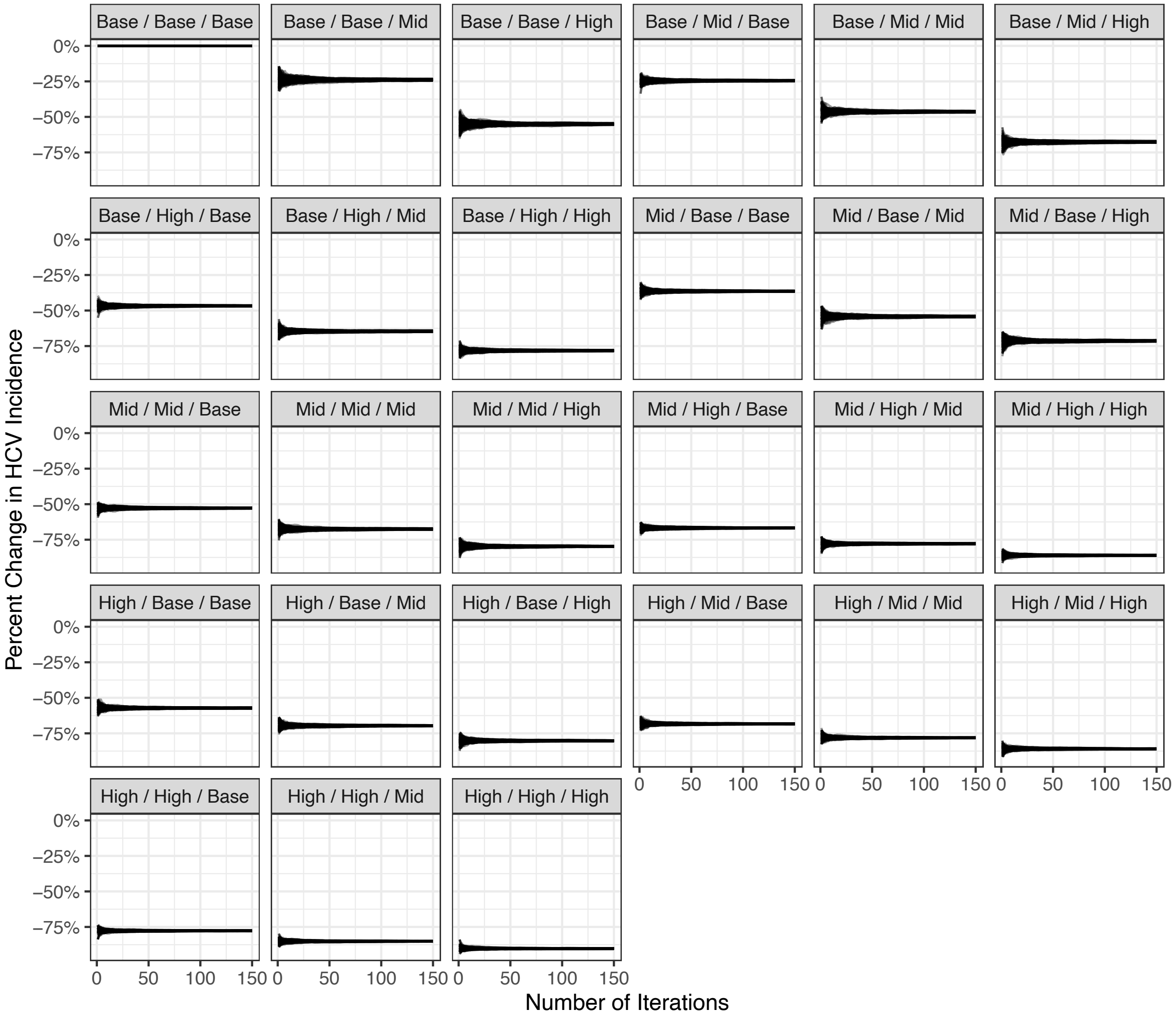

Footnote: Panel labels report cessation level first, prevention intervention level second, and test and treat level third. Baseline cessation rate is 16 per 100 person-years, moderate cessation rate is 31 per 100 person-years, and high cessation rate is 46 per 100 person-years. Baseline prevention intervention coverage for injection equipment sharing is 53%, moderate coverage is 68%, and high coverage is 83%. Baseline prevention intervention coverage for sex is 20%, moderate coverage is 35%, and high coverage is 50%. The percentage of people with HCV infection reaching SVR per year is 3% at baseline test and treat, 16% at moderate test and treat, and 28% at high test and treat. The percentage of people with HIV that are virally suppressed is 44% at baseline test and treat, 58% at moderate test and treat, and 71% at high test and treat.

#### Supplemental Results

*Supplemental Figure S14 Impacts of scaling single and combined interventions, including additional levels of test and treat*, on HIV and HCV infection incidence among people who inject drugs, compared to baseline.*

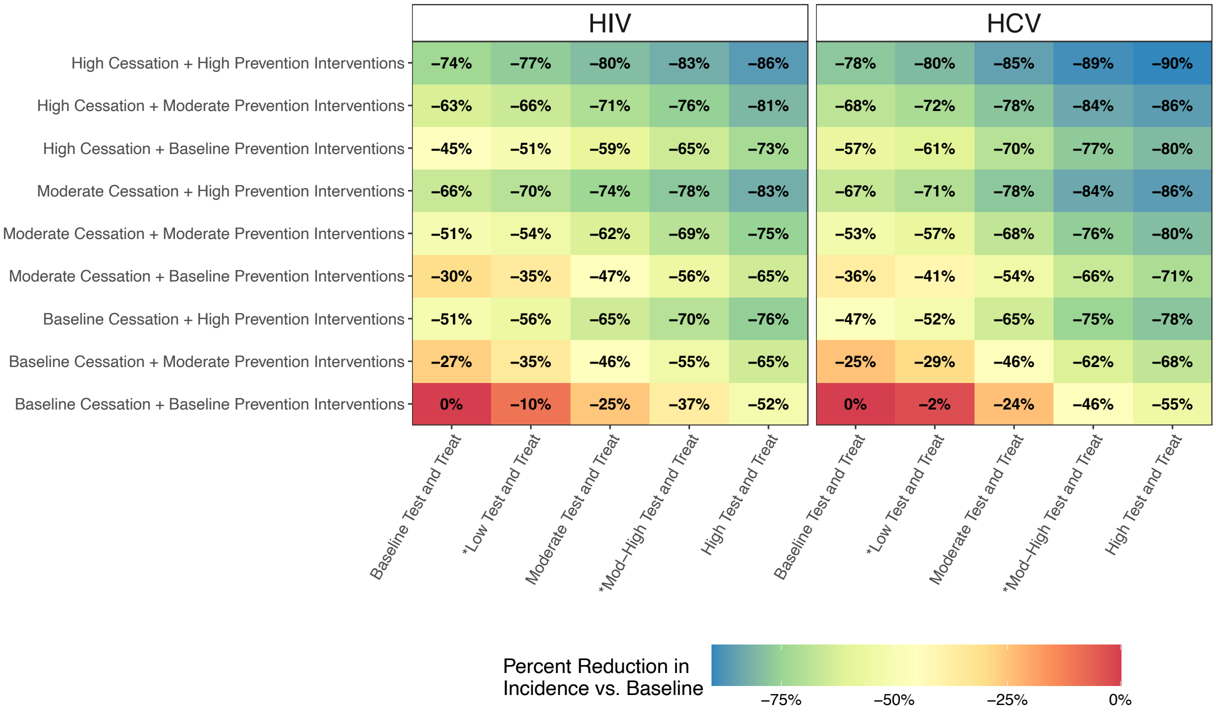

Footnote: Baseline cessation rate is 16 per 100 person-years, moderate cessation rate is 31 per 100 person-years, and high cessation rate is 46 per 100 person-years. Baseline prevention intervention coverage for injection equipment sharing is 53%, moderate coverage is 68%, and high coverage is 83%. Baseline prevention intervention coverage for sex is 20%, moderate coverage is 35%, and high coverage is 50%. The percentage of people with HCV infection reaching SVR per year is 3% at baseline test and treat, 7% at low test and treat, 16% at moderate test and treat, 24% at mod-high test and treat, and 28% at high test and treat. The percentage of people with HIV that are virally suppressed is 44% at baseline test and treat, 50% at low test and treat, 58% at moderate test and treat, 64% at mod-high test and treat, and 71% at high test and treat.

#### CHEERS Checklist

| Topic | No. | Item | Page where item is reported |
|---|---|---|---|
| **Title** | | | |
| | 1 | Identify the study as an economic evaluation and specify the interventions being compared. | N/A |
| **Abstract** | | | |
| | 2 | Provide a structured summary that highlights context, key methods, results, and alternative analyses. | 2-3 |
| **Introduction** | | | |
| **Background and objectives** | 3 | Give the context for the study, the study question, and its practical relevance for decision making in policy or practice. | 4-5 |
| **Methods** | | | |
| **Health economic analysis plan** | 4 | Indicate whether a health economic analysis plan was developed and where available. | N/A |
| **Study population** | 5 | Describe characteristics of the study population (such as age range, demographics, socioeconomic, or clinical characteristics). | 8 |
| **Setting and location** | 6 | Provide relevant contextual information that may influence findings. | 8; 24; 25 |
| **Comparators** | 7 | Describe the interventions or strategies being compared and why chosen. | 14-15 |
| **Perspective** | 8 | State the perspective(s) adopted by the study and why chosen. | N/A |
| **Time horizon** | 9 | State the time horizon for the study and why appropriate. | 15 |
| **Discount rate** | 10 | Report the discount rate(s) and reason chosen. | 15 |

| Topic | No. | Item | Page where item is reported |
|---|---|---|---|
| **Selection of outcomes** | 11 | Describe what outcomes were used as the measure(s) of benefit(s) and harm(s). | 15 |
| **Measurement of outcomes** | 12 | Describe how outcomes used to capture benefit(s) and harm(s) were measured. | 15-16 |
| **Valuation of outcomes** | 13 | Describe the population and methods used to measure and value outcomes. | 15-16; Appendix |
| **Measurement and valuation of resources and costs** | 14 | Describe how costs were valued. | N/A |
| **Currency, price date, and conversion** | 15 | Report the dates of the estimated resource quantities and unit costs, plus the currency and year of conversion. | N/A |
| **Rationale and description of model** | 16 | If modelling is used, describe in detail and why used. Report if the model is publicly available and where it can be accessed. | 5-6; 13; Appendix |
| **Analytics and assumptions** | 17 | Describe any methods for analysing or statistically transforming data, any extrapolation methods, and approaches for validating any model used. | 13; Appendix |
| **Characterising heterogeneity** | 18 | Describe any methods used for estimating how the results of the study vary for subgroups. | 25 |
| **Characterising distributional effects** | 19 | Describe how impacts are distributed across different individuals or adjustments made to reflect priority populations. | 25 |
| **Characterising uncertainty** | 20 | Describe methods to characterise any sources of uncertainty in the analysis. | 16; 25; Appendix |
| **Approach to engagement with patients and others affected by the study** | 21 | Describe any approaches to engage patients or service recipients, the general public, communities, or stakeholders (such as clinicians or payers) in the design of the study. | Not included beyond perspectives on author list. |
| **Results** | | | |

| Topic | No. | Item | Page where item is reported |
|---|---|---|---|
| **Study parameters** | 22 | Report all analytic inputs (such as values, ranges, references) including uncertainty or distributional assumptions. | Table 1; 16; 25 |
| **Summary of main results** | 23 | Report the mean values for the main categories of costs and outcomes of interest and summarise them in the most appropriate overall measure. | 16-17 |
| **Effect of uncertainty** | 24 | Describe how uncertainty about analytic judgments, inputs, or projections affect findings. Report the effect of choice of discount rate and time horizon, if applicable. | 21 |
| **Effect of engagement with patients and others affected by the study** | 25 | Report on any difference patient/service recipient, general public, community, or stakeholder involvement made to the approach or findings of the study | Not included beyond perspectives on author list. |
| **Discussion** | | | |
| **Study findings, limitations, generalisability, and current knowledge** | 26 | Report key findings, limitations, ethical or equity considerations not captured, and how these could affect patients, policy, or practice. | 24-25 |
| **Other relevant information** | | | |
| **Source of funding** | 27 | Describe how the study was funded and any role of the funder in the identification, design, conduct, and reporting of the analysis | 3; 16 |
| **Conflicts of interest** | 28 | Report authors conflicts of interest according to journal or International Committee of Medical Journal Editors requirements. | Completed |

Note: Items listed as N/A are not relevant to this analysis.

*From:* Husereau D, Drummond M, Augustovski F, et al. Consolidated Health Economic Evaluation Reporting Standards 2022 (CHEERS 2022) Explanation and Elaboration: A Report of the ISPOR CHEERS II Good Practices Task Force. Value Health 2022;25. doi:10.1016/j.jval.2021.10.008

#### References

1. Kanny D, Broz D, Finlayson T, et al. A Key Comprehensive System for Biobehavioral Surveillance of Populations Disproportionately Affected by HIV (National HIV Behavioral Surveillance): Cross-sectional Survey Study. *JMIR Public Health Surveill* 2022; 8: e39053.

2. Centers for Disease Control and Prevention. *HIV Infection Risk, Prevention, and Testing Behaviors among Persons Who Inject Drugs—National HIV Behavioral Surveillance: Injection Drug Use, 23 U.S. Cities, 2018*. HIV Surveillance Special Report 24, https://stacks.cdc.gov/view/cdc/106349 (February 2020).

3. McLaughlin M, Amaya A, Klevens M, et al. A review of factors associated with age of first injection. *J Psychoactive Drugs* 2020; 52: 412–420.

4. Park D, Oh S, Cano M, et al. Trends and distinct profiles of persons who inject drugs in the United States, 2015–2019. *Prev Med* 2022; 164: 107289.

5. Bradley H, Hall EW, Asher A, et al. Estimated Number of People Who Inject Drugs in the United States. *Clin Infect Dis Off Publ Infect Dis Soc Am* 2023; 76: 96–102.

6. Hall EW, Sullivan PS, Bradley H. Estimated Number of Injection-Involved Overdose Deaths in US States From 2000 to 2020: Secondary Analysis of Surveillance Data. *JMIR Public Health Surveill* 2024; 10: e49527.

7. Shah NG, Galai N, Celentano DD, et al. Longitudinal predictors of injection cessation and subsequent relapse among a cohort of injection drug users in Baltimore, MD, 1988-2000. *Drug Alcohol Depend* 2006; 83: 147–156.

8. Evans JL, Hahn JA, Lum PJ, et al. Predictors of injection drug use cessation and relapse in a prospective cohort of young injection drug users in San Francisco, CA (UFO Study). *Drug Alcohol Depend* 2009; 101: 152–157.

9. Dong H, Hayashi K, Singer J, et al. Trajectories of injection drug use among people who use drugs in Vancouver, Canada, 1996–2017: growth mixture modeling using data from prospective cohort studies. *Addiction* 2019; 114: 2173–2186.

10. Evans E, Li L, Min J, et al. Mortality among individuals accessing pharmacological treatment for opioid dependence in California, 2006-10. *Addict Abingdon Engl* 2015; 110: 996–1005.

11. Larney S, Tran LT, Leung J, et al. All-Cause and Cause-Specific Mortality Among People Using Extramedical Opioids: A Systematic Review and Meta-analysis. *JAMA Psychiatry* 2020; 77: 493–502.

12. Cepeda JA, Astemborski J, Kirk GD, et al. Rising role of prescription drugs as a portal to injection drug use and associated mortality in Baltimore, Maryland. *PLOS ONE* 2019; 14: e0213357.

13. Arias E, Xu JQ. *United States Life Tables, 2019*. Volume 70 Number 19, National Center for Health Statistics, https://www.cdc.gov/nchs/data/nvsr/nvsr70/nvsr70-19.pdf (2022).

14. Wang PL, Djerboua M, Flemming JA. Cause-specific mortality among patients with cirrhosis in a population-based cohort study in Ontario (2000–2017). *Hepatol Commun* 2023; 7: e00194.

15. van der Meer AJ, Veldt BJ, Feld JJ, et al. Association between sustained virological response and all-cause mortality among patients with chronic hepatitis C and advanced hepatic fibrosis. *JAMA* 2012; 308: 2584–2593.

16. Glaubius R, Kothegal N, Birhanu S, et al. Disease progression and mortality with untreated HIV infection: evidence synthesis of HIV seroconverter cohorts, antiretroviral treatment clinical cohorts and population-based survey data. *J Int AIDS Soc* 2021; 24: e25784.

17. Smith DJ, Combellick J, Jordan AE, et al. Hepatitis C virus (HCV) disease progression in people who inject drugs (PWID): A systematic review and meta-analysis. *Int J Drug Policy* 2015; 26: 911–921.

18. Erman A, Krahn MD, Hansen T, et al. Estimation of fibrosis progression rates for chronic hepatitis C: a systematic review and meta-analysis update. *BMJ Open* 2019; 9: e027491.

19. Guidelines for the Use of Antiretroviral Agents in Adults and Adolescents With HIV, https://clinicalinfo.hiv.gov/en/guidelines/hiv-clinical-guidelines-adult-and-adolescent-arv/coinfections-hepatitis-c-virus-hcv (2023, accessed 29 June 2024).

20. Thein H-H, Yi Q, Dore GJ, et al. Natural history of hepatitis C virus infection in HIV-infected individuals and the impact of HIV in the era of highly active antiretroviral therapy: a meta-analysis. *AIDS Lond Engl* 2008; 22: 1979–1991.

21. Blake A, Smith JE. Modeling Hepatitis C Elimination Among People Who Inject Drugs in New Hampshire. *JAMA Netw Open* 2021; 4: e2119092.

22. Aponte-Meléndez Y, Mateu-Gelabert P, Eckhardt B, et al. Hepatitis C virus care cascade among people who inject drugs in Puerto Rico: Minimal HCV treatment and substantial barriers to HCV care. *Drug Alcohol Depend Rep* 2023; 8: 100178.

23. Bull-Otterson L, Huang Y-LA, Zhu W, et al. Human Immunodeficiency Virus and Hepatitis C Virus Infection Testing Among Commercially Insured Persons Who Inject Drugs, United States, 2010-2017. *J Infect Dis* 2020; 222: 940–947.

24. Thompson WW, Symum H, Sandul A, et al. Vital Signs: Hepatitis C Treatment Among Insured Adults - United States, 2019-2020. *MMWR Morb Mortal Wkly Rep* 2022; 71: 1011–1017.

25. Kapadia SN, Zhang H, Gonzalez CJ, et al. Hepatitis C Treatment Initiation Among US Medicaid Enrollees. *JAMA Netw Open* 2023; 6: e2327326.

26. Berg T, Naumann U, Stoehr A, et al. Real-world effectiveness and safety of glecaprevir/pibrentasvir for the treatment of chronic hepatitis C infection: data from the German Hepatitis C-Registry. *Aliment Pharmacol Ther* 2019; 49: 1052–1059.

27. Cornberg M, Stoehr A, Naumann U, et al. Real-World Safety, Effectiveness, and Patient-Reported Outcomes in Patients with Chronic Hepatitis C Virus Infection Treated with Glecaprevir/Pibrentasvir: Updated Data from the German Hepatitis C-Registry (DHC-R). *Viruses* 2022; 14: 1541.

28. Sowah LA, Smeaton L, Brates I, et al. Perspectives on Adherence From the ACTG 5360 MINMON Trial: A Minimum Monitoring Approach With 12 Weeks of Sofosbuvir/Velpatasvir in Chronic Hepatitis C Treatment. *Clin Infect Dis Off Publ Infect Dis Soc Am* 2023; 76: 1959–1968.

29. Cunningham EB, Amin J, Feld JJ, et al. Adherence to sofosbuvir and velpatasvir among people with chronic HCV infection and recent injection drug use: The SIMPLIFY study. *Int J Drug Policy* 2018; 62: 14–23.

30. Hajarizadeh B, Cunningham EB, Valerio H, et al. Hepatitis C reinfection after successful antiviral treatment among people who inject drugs: A meta-analysis. *J Hepatol* 2020; 72: 643–657.

31. Esmaeili A, Mirzazadeh A, Carter GM, et al. Higher incidence of HCV in females compared to males who inject drugs: A systematic review and meta-analysis. *J Viral Hepat* 2017; 24: 117–127.

32. Valencia J, Alvaro-Meca A, Troya J, et al. High rates of early HCV reinfection after DAA treatment in people with recent drug use attended at mobile harm reduction units. *Int J Drug Policy* 2019; 72: 181–188.

33. Caven M, Malaguti A, Robinson E, et al. Impact of Hepatitis C treatment on behavioural change in relation to drug use in people who inject drugs: A systematic review. *Int J Drug Policy* 2019; 72: 169–176.

34. Coyle C, Moorman AC, Bartholomew T, et al. The HCV care continuum: linkage to HCV care and treatment among patients at an urban health network, Philadelphia, PA. *Hepatol Baltim Md* 2019; 70: 476–486.

35. Nosyk B, Lourenço L, Min JE, et al. Characterizing retention in HAART as a recurrent event process: insights into 'cascade churn'. *AIDS Lond Engl* 2015; 29: 1681–1689.

36. Saeed YA, Phoon A, Bielecki JM, et al. A Systematic Review and Meta-Analysis of Health Utilities in Patients With Chronic Hepatitis C. *Value Health J Int Soc Pharmacoeconomics Outcomes Res* 2020; 23: 127–137.

37. Tran BX, Nguyen LH, Ohinmaa A, et al. Longitudinal and cross sectional assessments of health utility in adults with HIV/AIDS: a systematic review and meta-analysis. *BMC Health Serv Res* 2015; 15: 7.

38. Nosyk B, Min JE, Krebs E, et al. The Cost-Effectiveness of Human Immunodeficiency Virus Testing and Treatment Engagement Initiatives in British Columbia, Canada: 2011–2013. *Clin Infect Dis* 2018; 66: 765–777.

39. Aden B, Dunning A, Nosyk B, et al. Impact of Illicit Drug Use on Health-Related Quality of Life in Opioid Dependent Patients Undergoing HIV Treatment. *J Acquir Immune Defic Syndr 1999* 2015; 70: 304–310.

40. Bernard CL, Owens DK, Goldhaber-Fiebert JD, et al. Estimation of the cost-effectiveness of HIV prevention portfolios for people who inject drugs in the United States: A model-based analysis. *PLoS Med* 2017; 14: e1002312.

41. Krivitsky PN, Handcock MS, Hunter DR, et al. statnet: Software tools for the Statistical Modeling of Network Data, http://statnet.org (2003).

42. Zhu L, Thompson WW, Hagan L, et al. Potential impact of curative and preventive interventions toward hepatitis C elimination in people who inject drugs–A network modeling study. *Int J Drug Policy* 2024; 130: 104539.

43. Chapin-Bardales J, Asher A, Broz D, et al. Hepatitis C virus infection and co-infection with HIV among persons who inject drugs in 10 U.S. cities—National HIV Behavioral Surveillance, 2018. *Int J Drug Policy* 2024; 104387.

44. Alter MJ. Epidemiology of viral hepatitis and HIV co-infection. *J Hepatol* 2006; 44: S6–S9.

45. Leyva Y, Page K, Shiboski S, et al. Per-Contact Infectivity of Hepatitis C Virus Acquisition in Association With Receptive Needle Sharing Exposures in a Prospective Cohort of Young Adult People who Inject Drugs in San Francisco, California. *Open Forum Infect Dis* 2020; 7: ofaa092.

46. Bartholomew TS, Onugha J, Bullock C, et al. Baseline prevalence and correlates of HIV and HCV infection among people who inject drugs accessing a syringe services program; Miami, FL. *Harm Reduct J* 2020; 17: 40.

47. Stout NK, Goldie SJ. Keeping the Noise Down: Common Random Numbers for Disease Simulation Modeling. *Health Care Manag Sci* 2008; 11: 399–406.
